# Sexual Function and Clitoral Anatomy after Vaginal Surgery with and without Midurethral Sling

**DOI:** 10.64898/2026.04.20.26351291

**Authors:** Shaniel T. Bowen, Pamela A. Moalli, Rebecca G. Rogers, Marlene M. Corton, Uduak U. Andy, Charles R. Rardin, Michael E. Hahn, Alison C. Weidner, David R. Ellington, Donna Mazloomdoost, Amaanti Sridhar, Marie G. Gantz, the NICHD Pelvic Floor Disorders Network

## Abstract

**Importance:** Sexual dysfunction can occur after midurethral sling (MUS) and transvaginal prolapse surgery. It remains unclear whether these procedures impact the clitoris, despite its role in sexual function and proximity to the MUS and vagina.

**Objectives:** To compare postoperative sexual function and clitoral features by MUS and vaginal surgery approach after transvaginal prolapse repair with/without concomitant MUS.

**Design:** Cross-sectional ancillary study of magnetic resonance imaging (MRI) and sexual function data from the Defining Mechanisms of Anterior Vaginal Wall Descent study.

**Setting:** Eight clinical sites in the US Pelvic Floor Disorders Network.

Participants: 88 women with uterovaginal prolapse who underwent vaginal mesh hysteropexy or vaginal hysterectomy with uterosacral ligament suspension with/without MUS between 2013-2015. Data were analyzed between September 2021-June 2023.

**Exposures:** Between June 2014-May 2018, participants underwent pelvic MRI 30-42 months after surgery, or earlier if reoperation was desired. Sexual activity and function at baseline and 24-48-month follow-up were evaluated using the Pelvic Organ Prolapse/Incontinence Sexual Questionnaire, IUGA-Revised (PISQ-IR). Clitoral features were obtained from postoperative MRI-based 3-dimensional models.

**Main Outcomes and Measures:** PISQ-IR scores and clitoral features (size, position).

**Results:** Eighty-two women (median [range] age, 65 [47-79] years) were analyzed: 45 MUS (22 hysteropexy, 23 hysterectomy) and 37 No-MUS (19 hysteropexy, 18 hysterectomy).

Postoperatively, 25 MUS, 12 No-MUS, 20 hysteropexy, and 17 hysterectomy patients were sexually active (SA). Overall, within the MUS and vaginal surgery groups, sexual function remained unchanged or improved (most PISQ-IR change from baseline scores were ≥0) among SA and NSA women. Among SA women after surgery, the MUS group (vs No-MUS) had a poorer PISQ-IR arousal/orgasm (SA-AO) score (median, 3.5 vs 4.3; P=.02). The hysteropexy group (vs hysterectomy) had less improvement in PISQ-IR SA-AO score (median, 0.0 vs 0.3; P=.01). Women with MUS (vs without) had a smaller clitoral glans thickness (median, 9.0 mm vs 10.0 mm; P=.008) and clitoral body volume (median, 2783.5 mm^3^ vs 3587.4 mm^3^; P=.01).

**Conclusions and Relevance:** SA women with MUS (vs without) or hysteropexy (vs hysterectomy) experienced poorer postoperative sexual function. MUS was linked to a smaller clitoris. Future studies should explore surgery-induced changes in clitoral anatomy and sexual function.

**KEY POINTS:** *Question:* How do sexual function and clitoral anatomy differ by midurethral sling placement and vaginal surgery approach?

*Findings:* This cross-sectional study compared patient-reported sexual function outcomes and 30-42-month postoperative magnetic resonance imaging-based 3-dimensional clitoral models of 82 women after vaginal prolapse surgery with or without concomitant midurethral sling. Midurethral sling (vs no sling) and vaginal mesh hysteropexy (vs vaginal hysterectomy) were associated with poorer postoperative sexual function outcomes. Additionally, midurethral sling was associated with a smaller clitoral glans and body.

*Meaning:* Midurethral sling and vaginal mesh hysteropexy were associated with, and may adversely alter, postoperative sexual function and/or clitoral anatomy.

*VISUAL ABSTRACT/PROMOTIONAL IMAGE:* 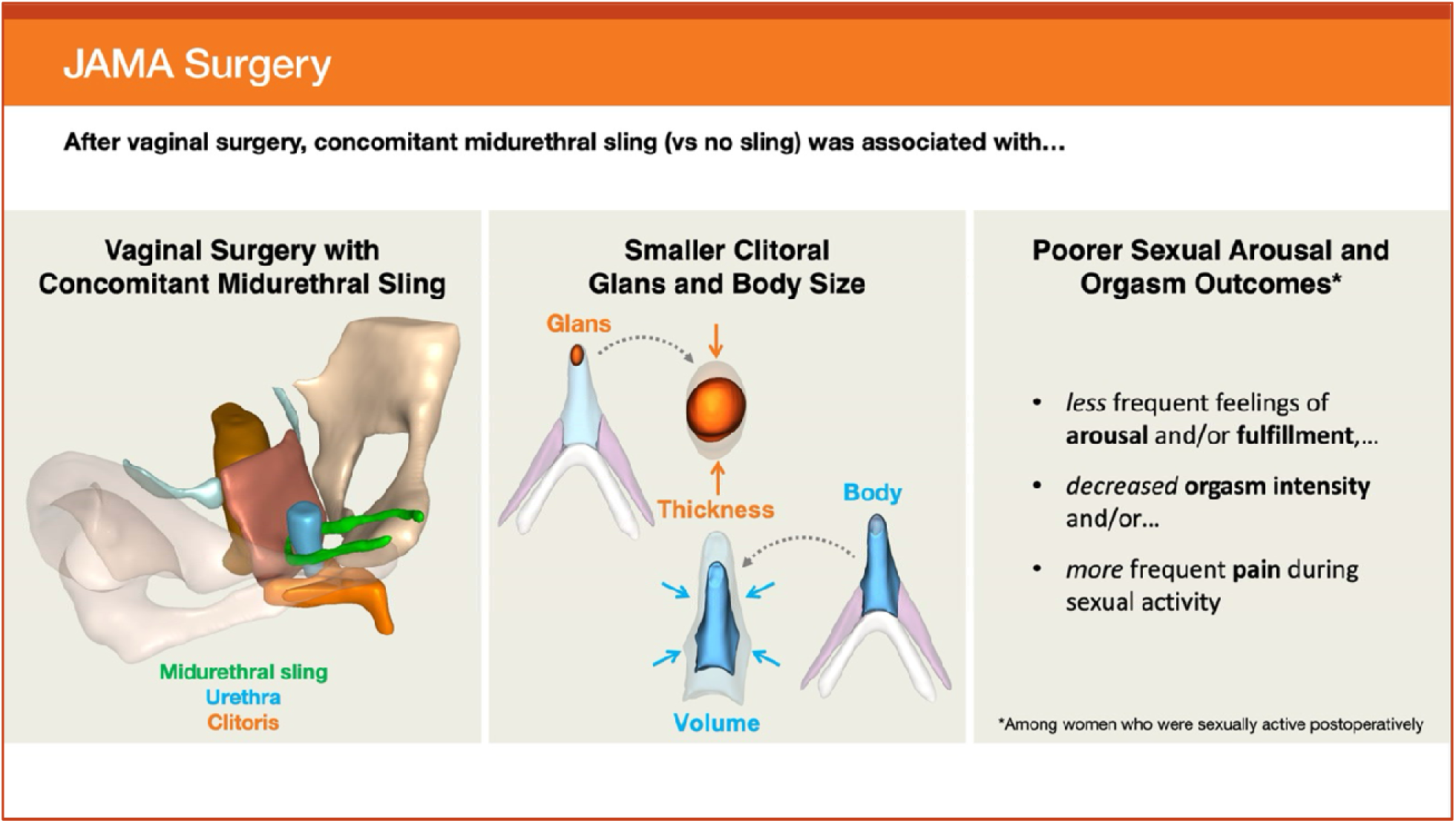

## INTRODUCTION

Transvaginal surgery with concomitant midurethral sling (MUS) is a common and often effective method to treat pelvic organ prolapse and stress urinary incontinence. Although the primary goal and measure of success for these procedures are the restoration of anatomical support and relief of prolapse and urinary symptoms,^1^ patients also expect their sexual function to improve or remain the same after surgery.^2^ While most women report better or unchanged sexual function following MUS and vaginal prolapse surgery,^3,4^ some women do experience postoperative sexual dysfunction. Meta-analyses have shown that the rate of sexual dysfunction after MUS placement can be as high as 20%,^5^ and about 10% after vaginal prolapse repair with or without mesh,^6^ including decreased arousal frequency, reduced orgasm intensity, and dyspareunia.^7^

It is suggested that intraoperative changes and injury to sexual anatomy and nearby structures within the surgical field can cause sexual dysfunction after MUS placement and transvaginal prolapse repair.^8^ Despite this, most studies on how MUS and vaginal surgery relate to sexual function have focused only on patient-reported outcomes. Therefore, little is known about the relationship between MUS, vaginal surgery, and the anatomy of the clitoral-vestibular bulb, despite the well-established link between the clitoris and sexual function.^9–11^.

To address this knowledge gap, sexual function and magnetic resonance imaging (MRI) data from the Defining Mechanisms of Anterior Vaginal Wall Descent (DEMAND) study^12,13^ were used to compare patient-reported sexual function and postoperative clitoral features by (1) MUS and (2) vaginal surgery approach among women who underwent transvaginal prolapse repair with and without concomitant MUS.

## METHODS

### Study Design

This was an ancillary analysis of the DEMAND study, a multicenter supplementary imaging study of a subgroup of women from the <u>S</u>tudy of <u>U</u>terine <u>P</u>rolapse Procedur<u>e</u>s-<u>R</u>andomized (SUPeR) trial.^13,14^ DEMAND described anatomical correlates and mechanisms of prolapse recurrence following treatment of uterovaginal prolapse using either vaginal mesh hysteropexy (hysteropexy) with Uphold LITE mesh (Boston Scientific) or vaginal hysterectomy with uterosacral ligament suspension (hysterectomy). The decision of MUS placement was based on the surgeon’s discretion. The study was conducted by the Pelvic Floor Disorders Network, received Institutional Review Board approval across all study sites, and obtained written informed consent from participants.

### Participants

Eighty-eight women recruited from SUPeR between June 2014-May 2018 were involved in the DEMAND study. Baseline (preoperative) and 24-48-month (on average) follow-up (postoperative) data obtained from SUPeR comprised demographics (including all self-reported races and ethnicities chosen from investigator-defined options for study inclusivity), clinical characteristics (medical history, Pelvic Organ Prolapse Quantification (POP-Q) measures), and patient-reported outcomes. Postoperative MRIs at 30-42 months (or before 30 months if reoperation was desired) were acquired from DEMAND. Detailed inclusion and exclusion criteria are provided in **eTable 1** in **Supplement 1**.

### Sexual Activity and Function Measures

Baseline (preoperative) and follow-up (SUPeR visits at 6, 12, 18, 24, 36, and 48 months postoperatively) sexual activity and function were evaluated using the Pelvic Organ Prolapse/Incontinence Sexual Questionnaire, IUGA Revised (PISQ-IR).^15,16^ For participants imaged near 30 and 42 months postoperatively, data were imputed from the next closest SUPeR visit to their MRI exam date with PISQ-IR data available (e.g., 24-, 36-, or 48-month follow-up visit).

Sexually active (SA) and not sexually active (NSA) groups were defined by PISQ-IR item 1, which asked if participants were sexually active with or without a partner. ^15,16^ For SA women, sexual function measures comprised dyspareunia (pain experienced sometimes, usually, or always during intercourse, based on PISQ-IR item 11), de novo dyspareunia (the absence of dyspareunia at baseline and the occurrence of dyspareunia at follow-up), PISQ-IR SA summary score, and subscale scores (Arousal-Orgasm (SA-AO), Condition Specific (SA-CS), and Condition Impact (SA-CI)).^15^ For NSA women, sexual function measures included dyspareunia (agreement with pain as a reason for avoiding sex, based on PISQ-IR item 2e) and PISQ-IR subscale scores (Condition Specific (NSA-CS) and Condition Impact (NSA-CI)).^15^ Higher PISQ-IR scores indicated better sexual function among SA women and a greater impact on sexual inactivity among NSA women.^16^

### Imaging Protocol

The MRI protocol has been published.^12^ In summary, postoperative axial T2-weighted MRIs were collected with 3-T scanners. Participants were imaged with a pelvic phased-array coil in the supine position at rest, with intravaginal contrast. Images were co-registered in 3D Slicer (v4.10.0, www.slicer.org)^17^ by using a 3D pelvic coordinate system defined by the pubic symphysis and ischial spines, where the midsagittal plane was given by its anterior-posterior (Y) and superior-inferior (Z) axes.^18^

### Image Segmentation and 3D Modeling

Manual segmentations of the clitoral-vestibular bulb complex (clitoris [glans, body, crura], vestibular bulbs), vagina, and urethra were performed in Slicer as described in previous studies.^10,19,20^ Briefly, the angle formed by (1) the ascending aspect of the clitoral body and (2) the medial aspect of the crura delineated the body from each crus. The posterior edge of the clitoral body was demarcated when only the body and crura were seen and extrapolated superiorly-inferiorly to separate the body and vestibular bulbs. The inferior edge of the clitoral body was marked where its midline septum last appeared prior to extending into the proximal glans.

Slice segmentations were then overlaid to construct 3D surface models, which were later smoothed in Blender (v2.83.3, Blender Foundation, Amsterdam, Netherlands) using a smoothing algorithm that preserved their original volume and shape while minimizing model-based measurement error.^19^ **Figure 1** shows an example of a final 3D model, and an accompanying MRI view, of a participant who underwent vaginal surgery with MUS.

**Figure 1.**
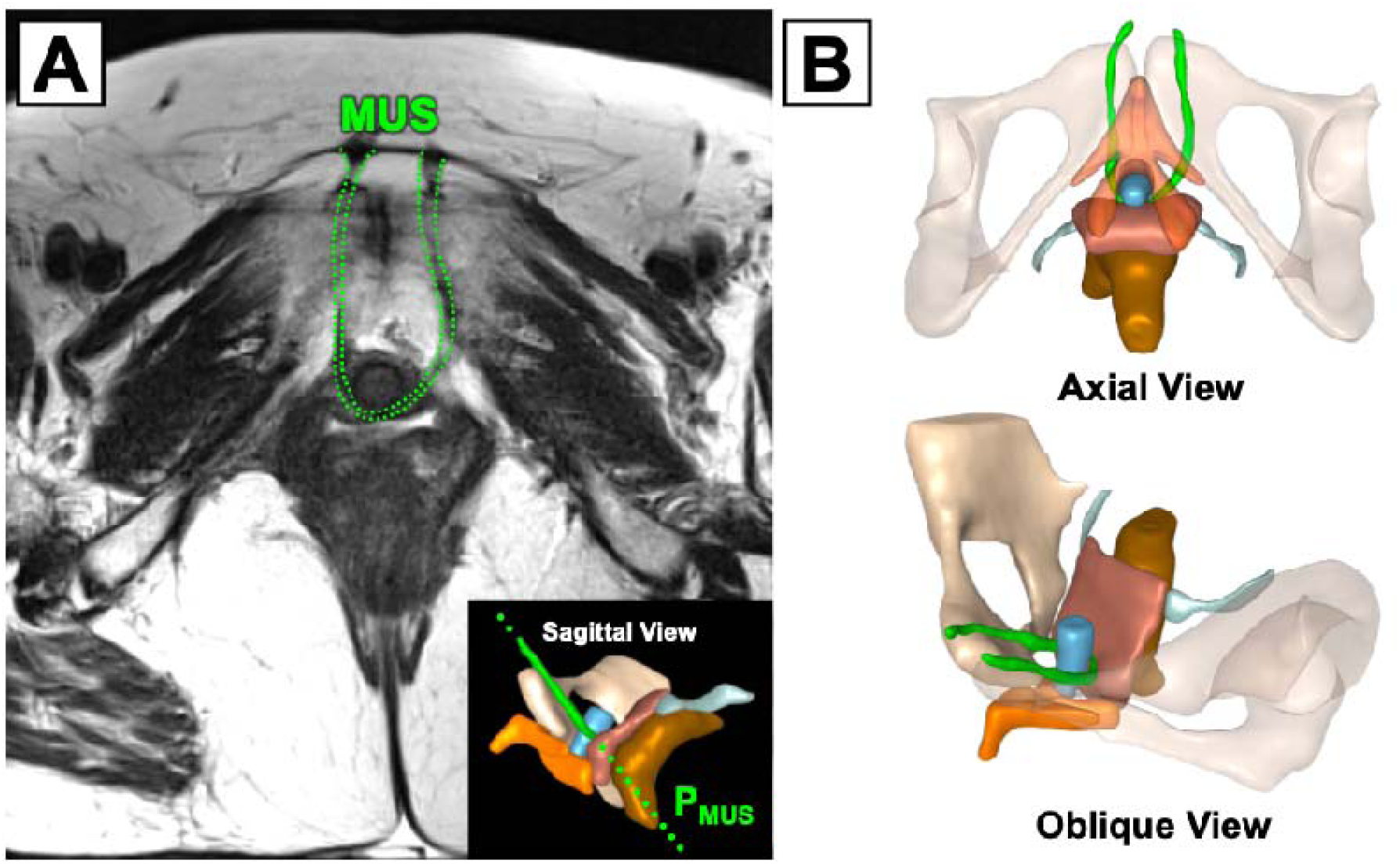
Visualization of a Vaginal Surgery with Concomitant Midurethral Sling. (**A**) Oblique view of an axial T2-weighted magnetic resonance image in the plane of the midurethral sling (P_MUS_, green dotted line) of a participant who underwent vaginal hysterectomy with uterosacral ligament suspension and concomitant MUS (green dotted contours). (**B**) Axial (top) and oblique (bottom) views of a 3-dimensional pelvic model of the same participant. Note the U-shaped aspect of the MUS (green) behind the distal urethra (blue) and lateral to the paraurethral glands, where each arm extends anteriorly into the retropubic space (behind the pubic bone) and inserts into the lower abdominal wall (∼2 cm from the midline). Also appreciate the proximity of the MUS relative to the clitoral-vestibular bulb complex (orange) and pubic symphysis (attachment site of clitoral neurovascular structures involved in the sexual response).

### Clitoral-Vestibular Bulb Measures

A Mathematica (v12.2.2.0, Wolfram Research, Champaign, IL, USA) custom algorithm was used to perform automated, model-based dimensional and position (pelvic location, urethrovaginal distance) measurements of the clitoral-vestibular bulb complex (**Figure 2**).^19^ Each clitoral substructure (glans, body, left and right crus) was fitted to a 3D bounding box whose dimensions defined its length, width, and thickness. The volumes of the clitoral-vestibular bulb complex and its individual components were derived from the volume enclosed by their respective surface models.

**Figure 2.**
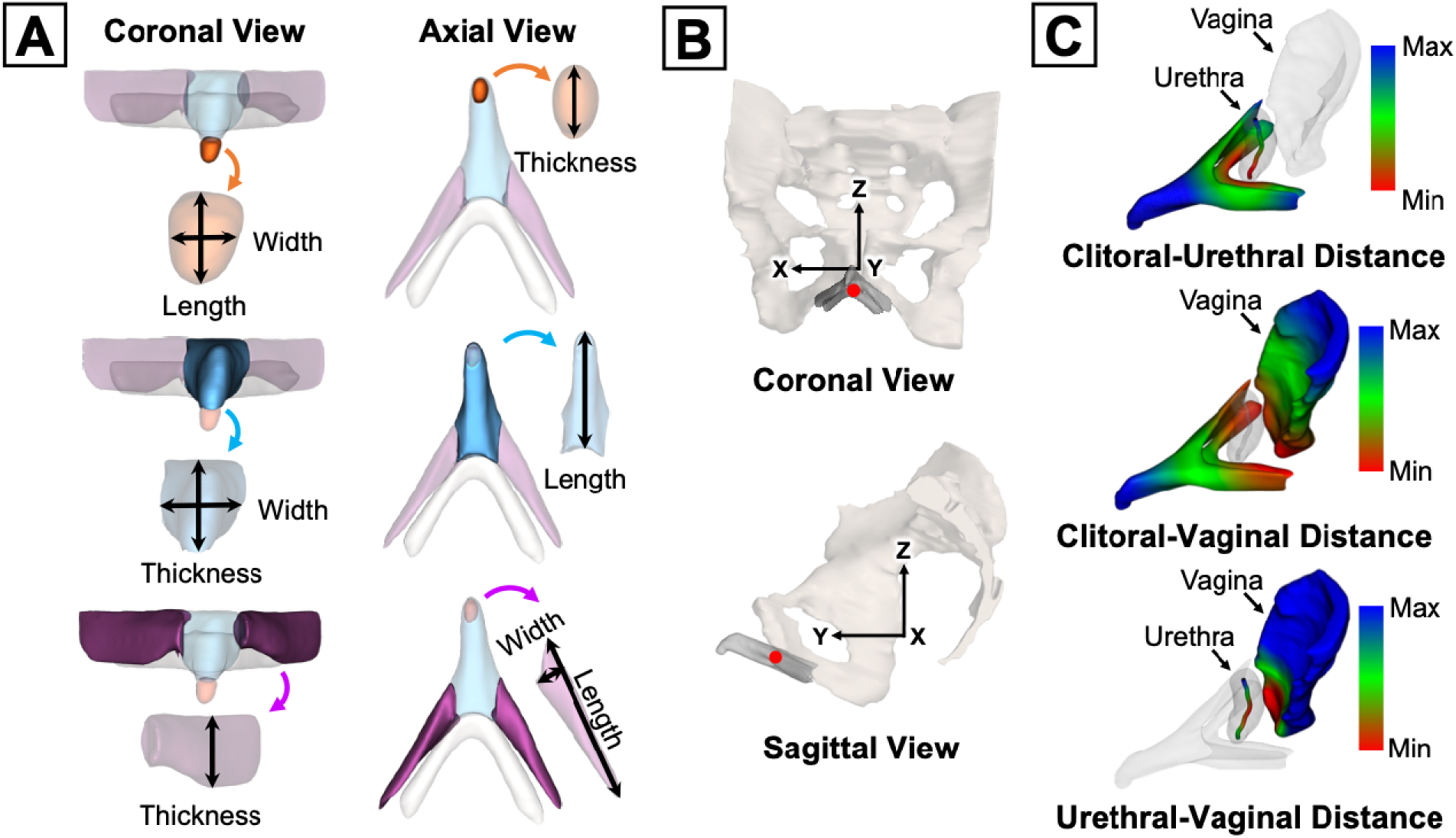
Clitoral-Vestibular Bulb Anatomy Measurements. (**A**) Coronal and axial views of dimension measurements of the clitoral glans (orange), body (light blue), and crura (purple). Volume measurements of the clitoral-vestibular bulb complex and the vestibular bulbs (white) are defined as the volumes within their respective 3-dimensional surface models. (**B**) Coronal and sagittal views of the clitoral-vestibular bulb complex position (red point) given by the centroid of the 3-dimensional model relative to the pelvic coordinate system (black arrows), where the *X*, *Y*, and *Z* axes define the medial-lateral, anterior-posterior, and superior-inferior position. (**C**) Distance measurements between the clitoris, urethra (urethral centerline), and vagina (anterior vaginal wall) were calculated from the minimum surface-to-surface distances (red areas) between structures of interest.

The position of the clitoral-vestibular bulb complex relative to the pelvis via the pelvic coordinate system was computed from its centroid, where its X, Y, and Z coordinates corresponded to its medial-lateral, anterior-posterior, and superior-inferior position. The position of the clitoral-vestibular bulb complex with respect to the urethra and vagina was quantified by its minimum surface-to-surface distance between the urethral centerline and the anterior vaginal wall, respectively.

### Outcomes

Outcome measures included PISQ-IR scores (summary, subscale, item) and clitoral-vestibular bulb dimensions and position (pelvic, urethrovaginal distances).

### Statistical Methods

Based on the primary DEMAND study, a sample size of 40 per vaginal surgery group was estimated to detect a moderate effect size (0.5 standard deviation) in outcome measures with 80% power.^13^ Baseline and follow-up patient characteristics, sexual activity and function measures, and clitoral-vestibular bulb measures were described as counts (percentages) or medians (interquartile ranges) and compared by (1) MUS and (2) vaginal surgery type using Fisher’s exact tests for categorical variables and Wilcoxon Rank-Sum tests for continuous variables. Given the exploratory nature of the analysis, no multiple-comparison adjustments were made. Thus, results should be interpreted with appropriate caution.

## RESULTS

### Study Population

Of the 88 women from the primary DEMAND cohort, six were excluded due to poor delineation of the vaginal border on MRI, leaving 82 included in this ancillary analysis: 45 MUS (22 hysteropexy, 23 hysterectomy) and 37 No-MUS (19 hysteropexy, 18 hysterectomy), with each vaginal surgery group consisting of 41 women. Seventy-five women were imaged at 30-42 months, 6 prior to 30 months, and 1 at 48 months postoperatively (**eFigures 1-2** in **Supplement 1**). The median (range) age, body mass index (BMI), and vaginal parity were 65 (47-79) years, 28 (19-39) kg/m², and 3 (0-10). Most of the study population were White (82%), postmenopausal (98%), married/living with a partner (63%), and had stage III/IV POP (76%) at baseline.

Comparisons of baseline and follow-up patient characteristics by MUS and vaginal surgery type are shown in **Table 1** and **eTable 3** in **Supplement 1**, respectively. At baseline, women with MUS (vs without) had a lower weight (median, 68.0 kg vs 77.0 kg; difference, −8.0 kg [95%CI, −13.0 to −3.0]; P=.002), body mass index (median, 25.6 kg/m^2^ vs 29.8 kg/m^2^; difference, −3.1 kg/m^2^ [95%CI, −4.8 to −1.2];P=.002), rate of obesity (22% vs 46%; risk difference, −24% [95%CI, −44% to −3%]; P=.03), and rate of prior stress urinary incontinence surgery (0% vs 14%; P=.02). Postoperatively at 24-48 months, the hysteropexy group (vs hysterectomy) showed better anterior vaginal wall support (POP-Q Point Ba) (median, −1.5 cm vs −1.0 cm; difference, −1.0 cm [95%CI, −1.5 to 0.0]; P=.02), a longer total vaginal length (TVL) (median, 9.0 cm vs 8.0 cm; difference, 1.0 cm [95%CI, 0.0 to 1.0]; P=.002), and less TVL shortening (change from baseline) (median, −0.5 cm vs −1.0 cm; difference, 1.0 cm [95%CI, 0.0 to 1.3]; P=.009).

**Table 1.**
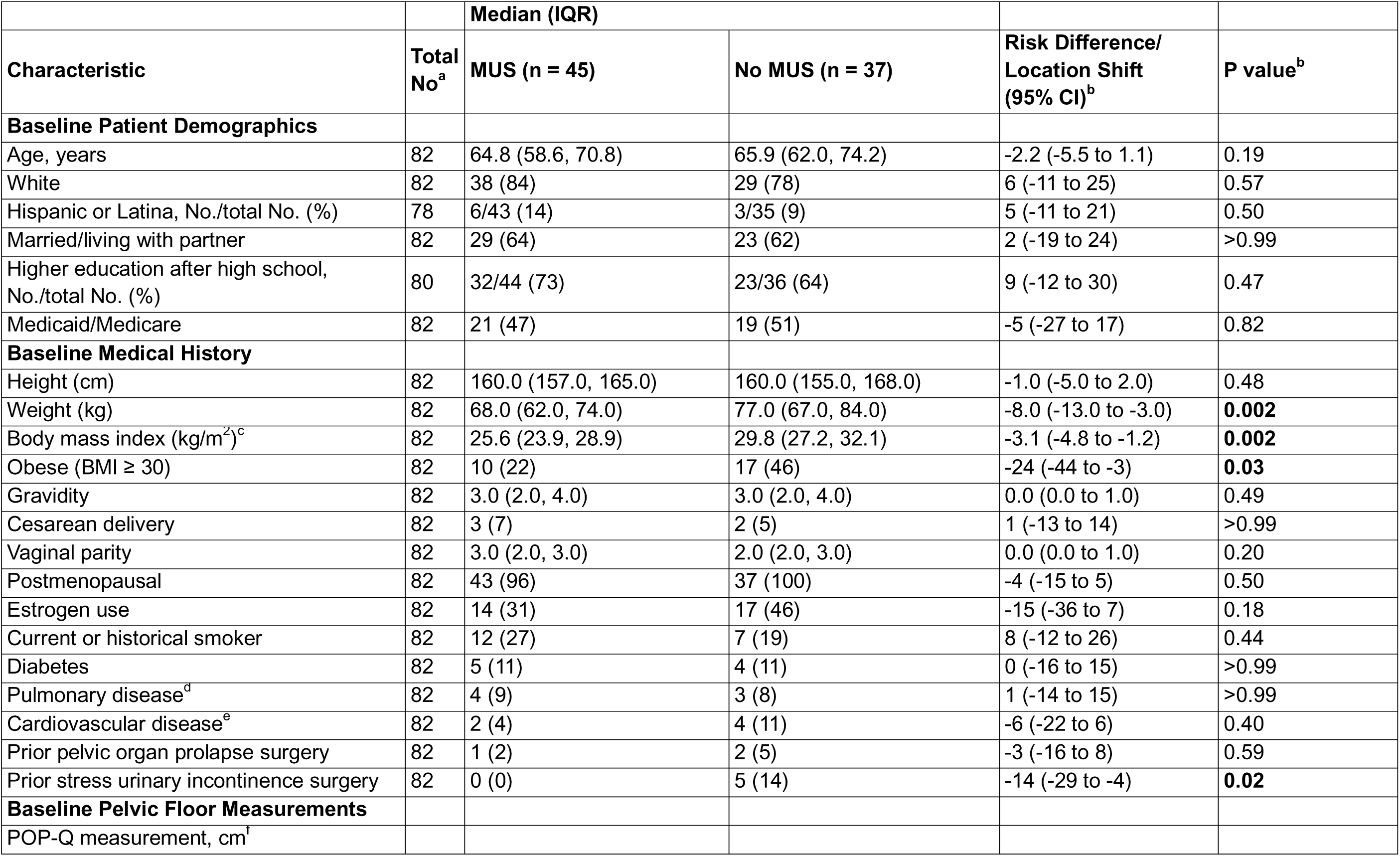
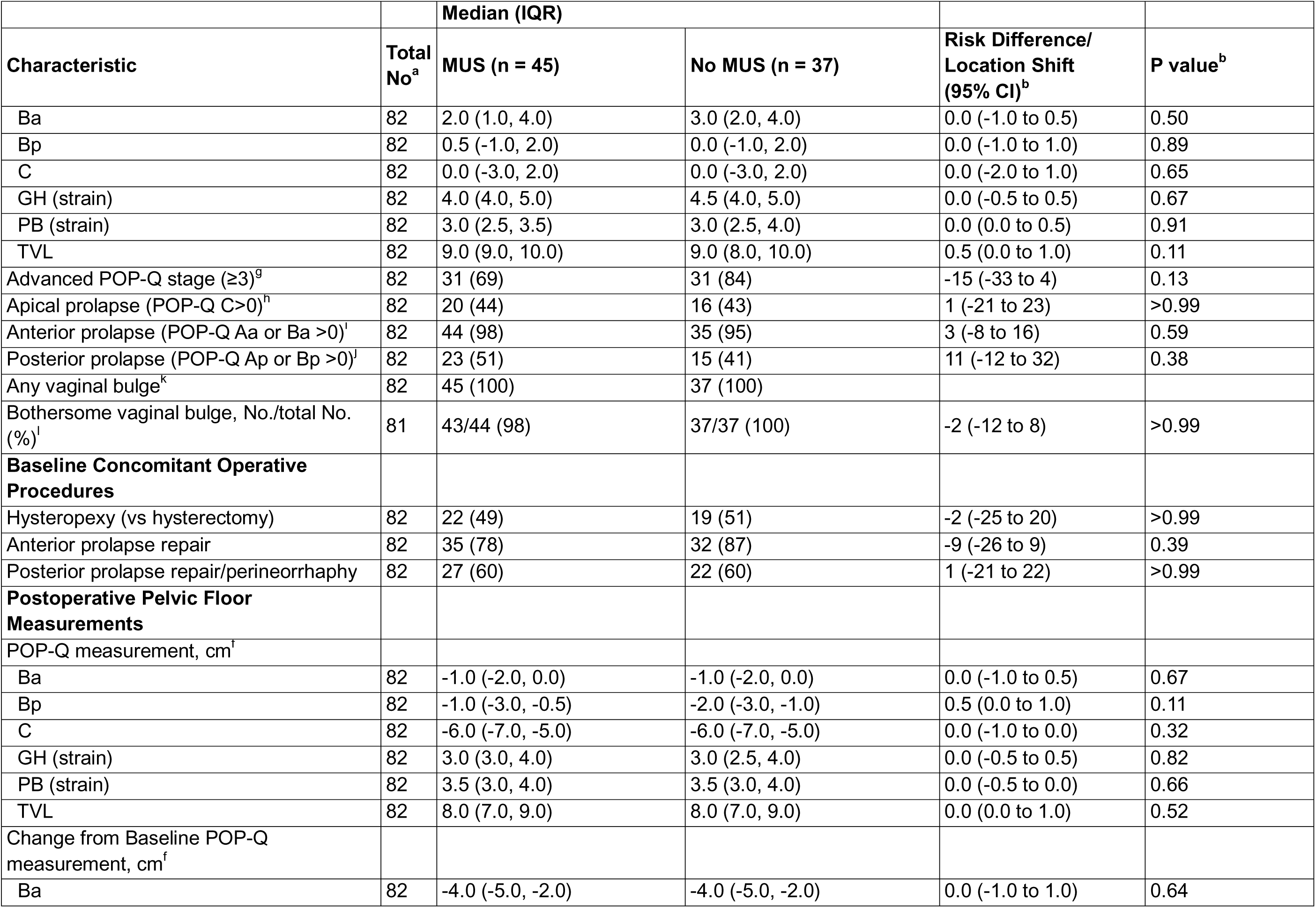
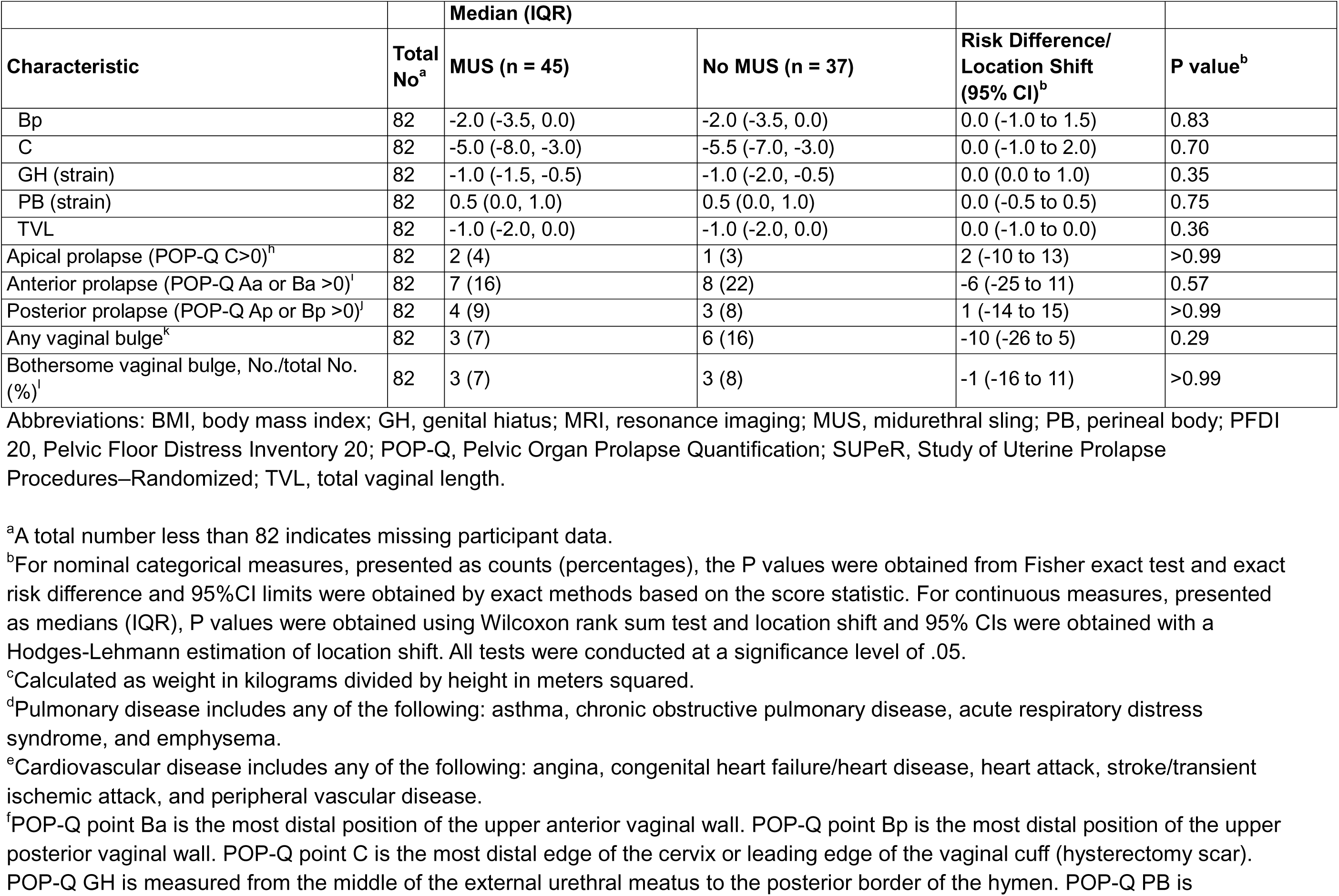
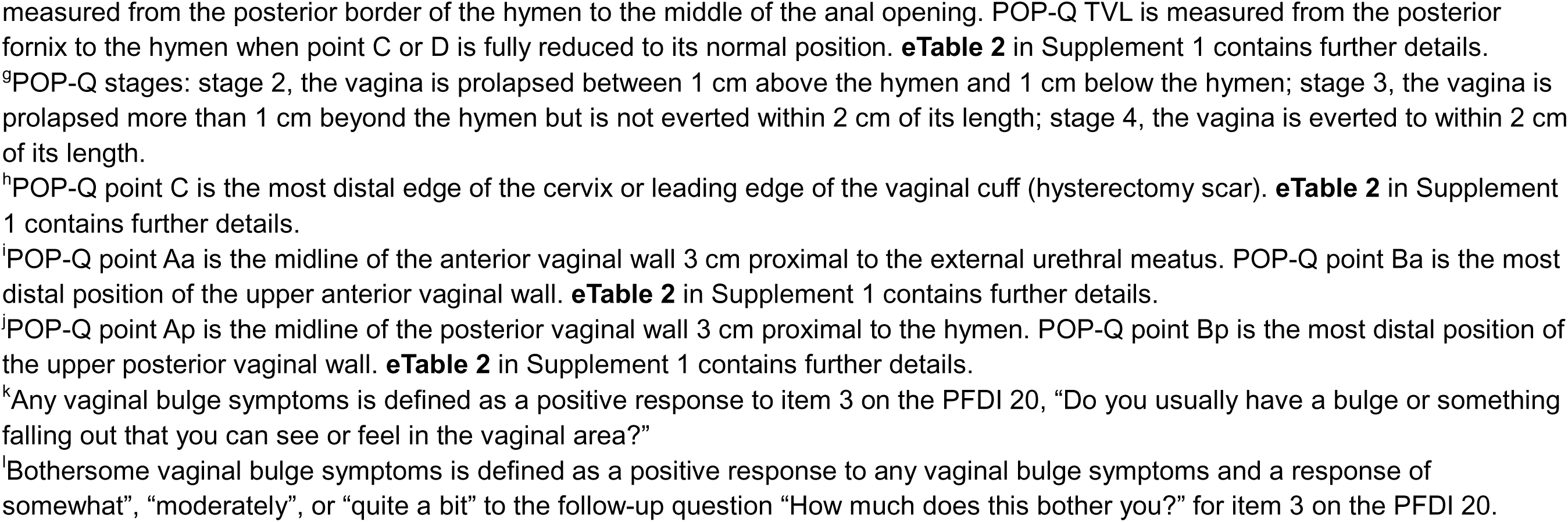
Baseline and Postoperative Demographic and Clinical Characteristics After Vaginal Prolapse Repair by Concomitant Midurethral Sling (MUS)

### Sexual Activity and Function

**Table 2** and **eTable 4** in **Supplement 1** show sexual activity and function outcomes before and after surgery stratified by MUS and vaginal surgery approach, respectively. At baseline, 37 (45%) women were SA and 45 (55%) were NSA, of whom 17 were SA with dyspareunia and 14 were NSA due to dyspareunia. Preoperative sexual activity status did not differ by MUS (P=.12) or vaginal surgery approach (P>.99). The rate of preoperative dyspareunia among SA and NSA women was similar between the MUS and vaginal surgery groups.

**Table 2.**
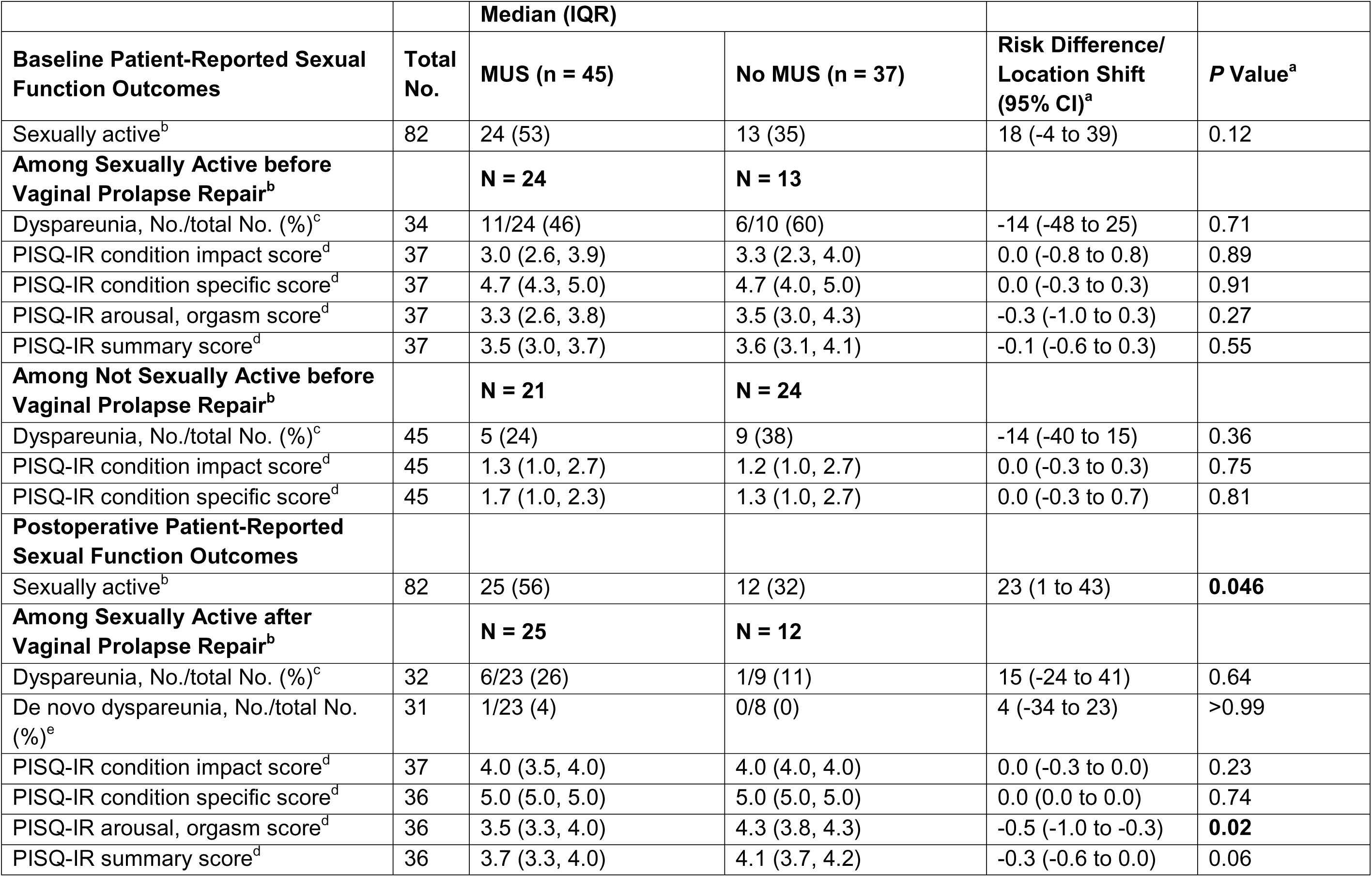
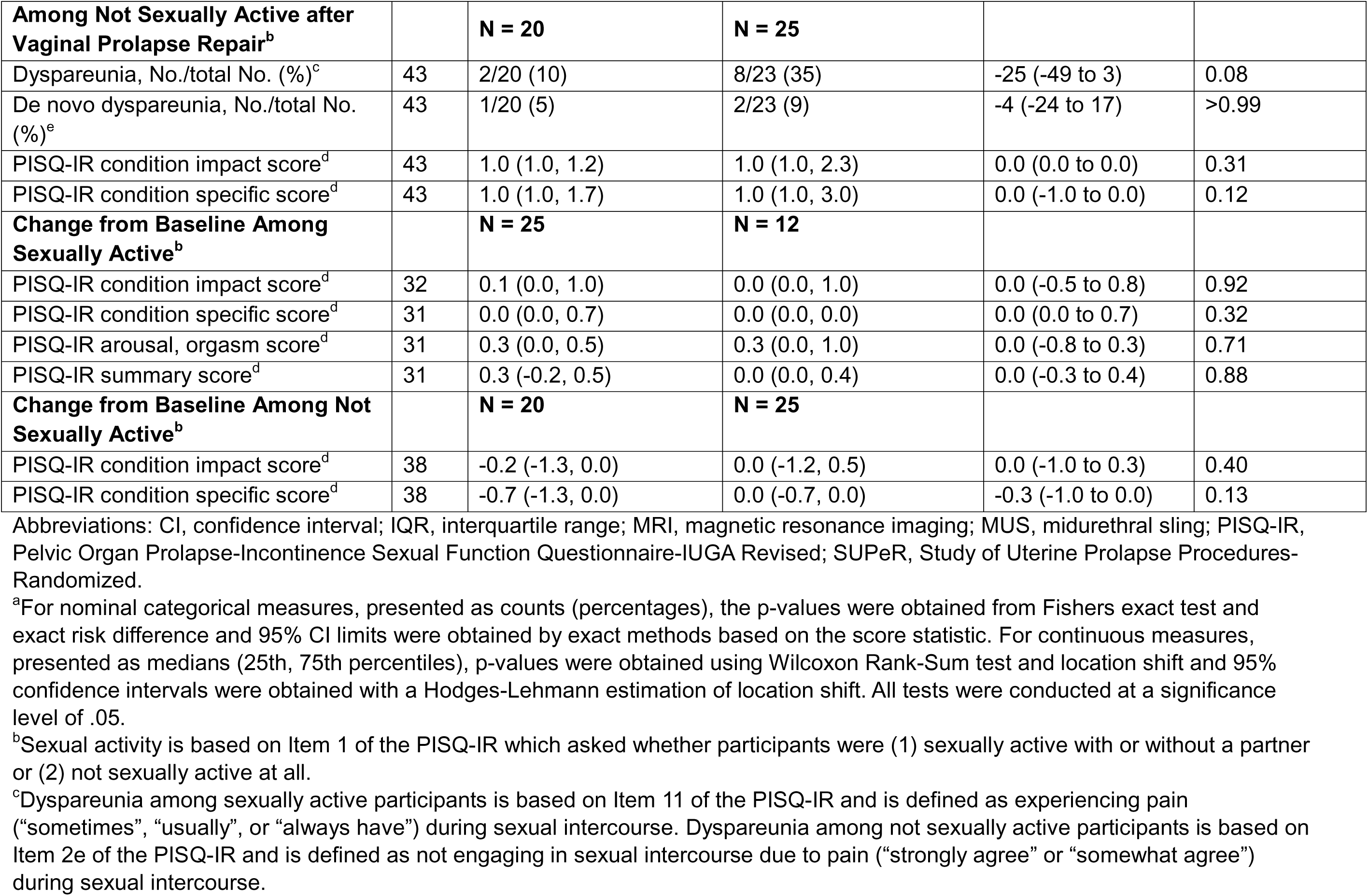
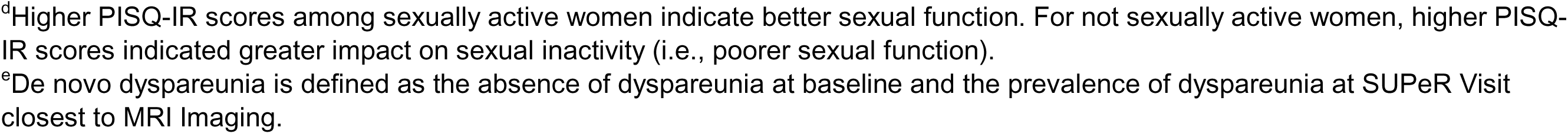
Baseline and Postoperative Sexual Function Outcomes After Vaginal Prolapse Repair by Concomitant Midurethral Sling (MUS)

|  |  | Median (IQR) |  |  |  |
| --- | --- | --- | --- | --- | --- |
| Baseline Patient-Reported Sexual Function Outcomes | Total No. | MUS (n = 45) | No MUS (n = 37) | Risk Difference/ Location Shift (95% CI)<sup>a</sup> | P Value<sup>a</sup> |
| Sexually active<sup>b</sup> | 82 | 24 (53) | 13 (35) | 18 (-4 to 39) | 0.12 |
| **Among Sexually Active before Vaginal Prolapse Repair<sup>b</sup>** |  | **N = 24** | **N = 13** |  |  |
| Dyspareunia, No./total No. (%)<sup>c</sup> | 34 | 11/24 (46) | 6/10 (60) | -14 (-48 to 25) | 0.71 |
| PISQ-IR condition impact score<sup>d</sup> | 37 | 3.0 (2.6, 3.9) | 3.3 (2.3, 4.0) | 0.0 (-0.8 to 0.8) | 0.89 |
| PISQ-IR condition specific score<sup>d</sup> | 37 | 4.7 (4.3, 5.0) | 4.7 (4.0, 5.0) | 0.0 (-0.3 to 0.3) | 0.91 |
| PISQ-IR arousal, orgasm score<sup>d</sup> | 37 | 3.3 (2.6, 3.8) | 3.5 (3.0, 4.3) | -0.3 (-1.0 to 0.3) | 0.27 |
| PISQ-IR summary score<sup>d</sup> | 37 | 3.5 (3.0, 3.7) | 3.6 (3.1, 4.1) | -0.1 (-0.6 to 0.3) | 0.55 |
| **Among Not Sexually Active before Vaginal Prolapse Repair<sup>b</sup>** |  | **N = 21** | **N = 24** |  |  |
| Dyspareunia, No./total No. (%)<sup>c</sup> | 45 | 5 (24) | 9 (38) | -14 (-40 to 15) | 0.36 |
| PISQ-IR condition impact score<sup>d</sup> | 45 | 1.3 (1.0, 2.7) | 1.2 (1.0, 2.7) | 0.0 (-0.3 to 0.3) | 0.75 |
| PISQ-IR condition specific score<sup>d</sup> | 45 | 1.7 (1.0, 2.3) | 1.3 (1.0, 2.7) | 0.0 (-0.3 to 0.7) | 0.81 |
| **Postoperative Patient-Reported Sexual Function Outcomes** |  |  |  |  |  |
| Sexually active<sup>b</sup> | 82 | 25 (56) | 12 (32) | 23 (1 to 43) | **0.046** |
| **Among Sexually Active after Vaginal Prolapse Repair<sup>b</sup>** |  | **N = 25** | **N = 12** |  |  |
| Dyspareunia, No./total No. (%)<sup>c</sup> | 32 | 6/23 (26) | 1/9 (11) | 15 (-24 to 41) | 0.64 |
| De novo dyspareunia, No./total No. (%)<sup>e</sup> | 31 | 1/23 (4) | 0/8 (0) | 4 (-34 to 23) | >0.99 |
| PISQ-IR condition impact score<sup>d</sup> | 37 | 4.0 (3.5, 4.0) | 4.0 (4.0, 4.0) | 0.0 (-0.3 to 0.0) | 0.23 |
| PISQ-IR condition specific score<sup>d</sup> | 36 | 5.0 (5.0, 5.0) | 5.0 (5.0, 5.0) | 0.0 (0.0 to 0.0) | 0.74 |
| PISQ-IR arousal, orgasm score<sup>d</sup> | 36 | 3.5 (3.3, 4.0) | 4.3 (3.8, 4.3) | -0.5 (-1.0 to -0.3) | **0.02** |
| PISQ-IR summary score<sup>d</sup> | 36 | 3.7 (3.3, 4.0) | 4.1 (3.7, 4.2) | -0.3 (-0.6 to 0.0) | 0.06 |

| **Among Not Sexually Active after Vaginal Prolapse Repair<sup>b</sup>** |  | **N = 20** | **N = 25** |  |  |
| --- | --- | --- | --- | --- | --- |
| Dyspareunia, No./total No. (%)<sup>c</sup> | 43 | 2/20 (10) | 8/23 (35) | -25 (-49 to 3) | 0.08 |
| De novo dyspareunia, No./total No. (%)<sup>e</sup> | 43 | 1/20 (5) | 2/23 (9) | -4 (-24 to 17) | >0.99 |
| PISQ-IR condition impact score<sup>d</sup> | 43 | 1.0 (1.0, 1.2) | 1.0 (1.0, 2.3) | 0.0 (0.0 to 0.0) | 0.31 |
| PISQ-IR condition specific score<sup>d</sup> | 43 | 1.0 (1.0, 1.7) | 1.0 (1.0, 3.0) | 0.0 (-1.0 to 0.0) | 0.12 |
| **Change from Baseline Among Sexually Active<sup>b</sup>** |  | **N = 25** | **N = 12** |  |  |
| PISQ-IR condition impact score<sup>d</sup> | 32 | 0.1 (0.0, 1.0) | 0.0 (0.0, 1.0) | 0.0 (-0.5 to 0.8) | 0.92 |
| PISQ-IR condition specific score<sup>d</sup> | 31 | 0.0 (0.0, 0.7) | 0.0 (0.0, 0.0) | 0.0 (0.0 to 0.7) | 0.32 |
| PISQ-IR arousal, orgasm score<sup>d</sup> | 31 | 0.3 (0.0, 0.5) | 0.3 (0.0, 1.0) | 0.0 (-0.8 to 0.3) | 0.71 |
| PISQ-IR summary score<sup>d</sup> | 31 | 0.3 (-0.2, 0.5) | 0.0 (0.0, 0.4) | 0.0 (-0.3 to 0.4) | 0.88 |
| **Change from Baseline Among Not Sexually Active<sup>b</sup>** |  | **N = 20** | **N = 25** |  |  |
| PISQ-IR condition impact score<sup>d</sup> | 38 | -0.2 (-1.3, 0.0) | 0.0 (-1.2, 0.5) | 0.0 (-1.0 to 0.3) | 0.40 |
| PISQ-IR condition specific score<sup>d</sup> | 38 | -0.7 (-1.3, 0.0) | 0.0 (-0.7, 0.0) | -0.3 (-1.0 to 0.0) | 0.13 |

Abbreviations: CI, confidence interval; IQR, interquartile range; MRI, magnetic resonance imaging; MUS, midurethral sling; PISQ-IR, Pelvic Organ Prolapse-Incontinence Sexual Function Questionnaire-IUGA Revised; SUPeR, Study of Uterine Prolapse Procedures-Randomized.
<sup>a</sup>For nominal categorical measures, presented as counts (percentages), the p-values were obtained from Fishers exact test and exact risk difference and 95% CI limits were obtained by exact methods based on the score statistic. For continuous measures, presented as medians (25th, 75th percentiles), p-values were obtained using Wilcoxon Rank-Sum test and location shift and 95% confidence intervals were obtained with a Hodges-Lehmann estimation of location shift. All tests were conducted at a significance level of .05.
<sup>b</sup>Sexual activity is based on Item 1 of the PISQ-IR which asked whether participants were (1) sexually active with or without a partner or (2) not sexually active at all.
<sup>c</sup>Dyspareunia among sexually active participants is based on Item 11 of the PISQ-IR and is defined as experiencing pain ("sometimes", "usually", or "always have") during sexual intercourse. Dyspareunia among not sexually active participants is based on Item 2e of the PISQ-IR and is defined as not engaging in sexual intercourse due to pain ("strongly agree" or "somewhat agree") during sexual intercourse.
<sup>d</sup>Higher PISQ-IR scores among sexually active women indicate better sexual function. For not sexually active women, higher PISQ- IR scores indicated greater impact on sexual inactivity (i.e., poorer sexual function). <sup>e</sup>De novo dyspareunia is defined as the absence of dyspareunia at baseline and the prevalence of dyspareunia at SUPeR Visit closest to MRI Imaging.

As noted in prior literature, patients may change sexual activity status over time.^3^ Postoperatively, 5 women (6%) became SA and 5 became NSA. Thus, 37 (45%) women were SA and 45 (55%) were NSA at follow-up, of whom 7 were SA with dyspareunia and 10 were NSA due to dyspareunia. De novo dyspareunia was reported by 1 SA woman and 3 NSA women. Postoperatively, more women were sexually active in the MUS group than the No MUS group (P=.046). Postoperative sexual activity status was similar among the vaginal surgery groups (P=.66). There were no differences in the rate of postoperative dyspareunia or de novo dyspareunia by MUS or vaginal surgery approach.

Regarding PISQ-IR outcomes, sexual function among SA and NSA women largely remained either unchanged or improved after surgery within the MUS and vaginal surgery groups, as indicated by most change in baseline PISQ-IR scores being greater than or equal to zero. However, among SA women after vaginal surgery, those with MUS (vs without) had a lower SA-AO subscale score (median, 3.5 vs 4.3; difference, −0.5 [95%CI, −1.0 to −0.3]; P=.02), indicating worse postoperative sexual function associated with less frequent feelings of arousal and/or fulfillment, decreased orgasm intensity, and/or more frequent pain during sexual activity. Additionally, among SA women, those who received hysteropexy (vs hysterectomy) showed less improvement (change from baseline) in SA-AO scores (median, 0.0 vs 0.3; difference, −0.5 [95%CI, −0.8 to 0.0]; P=.01).

### Clitoral-Vestibular Bulb Measures

Comparisons of postoperative measurements of the clitoral-vestibular bulb between the MUS groups and vaginal surgery groups are presented in Table 3 and eTable 5 in Supplement 1, respectively. Women with MUS (vs without) had a smaller clitoral glans thickness (median, 9.0 mm vs 10.0 mm; difference, −0.8 mm [95%CI, −1.5 to −0.2]; P=.008) and a smaller clitoral body volume (median, 2783.5 mm^3^ vs 3587.4 mm^3^; difference, −557.5 mm^3^ [95%CI, −952.7 to −130.6]; P=.01). The position of the clitoral-vestibular bulb was similar across MUS groups. There were no significant differences in the dimensions or position of the clitoral-vestibular bulb based on vaginal surgery approach.

**Table 3.**
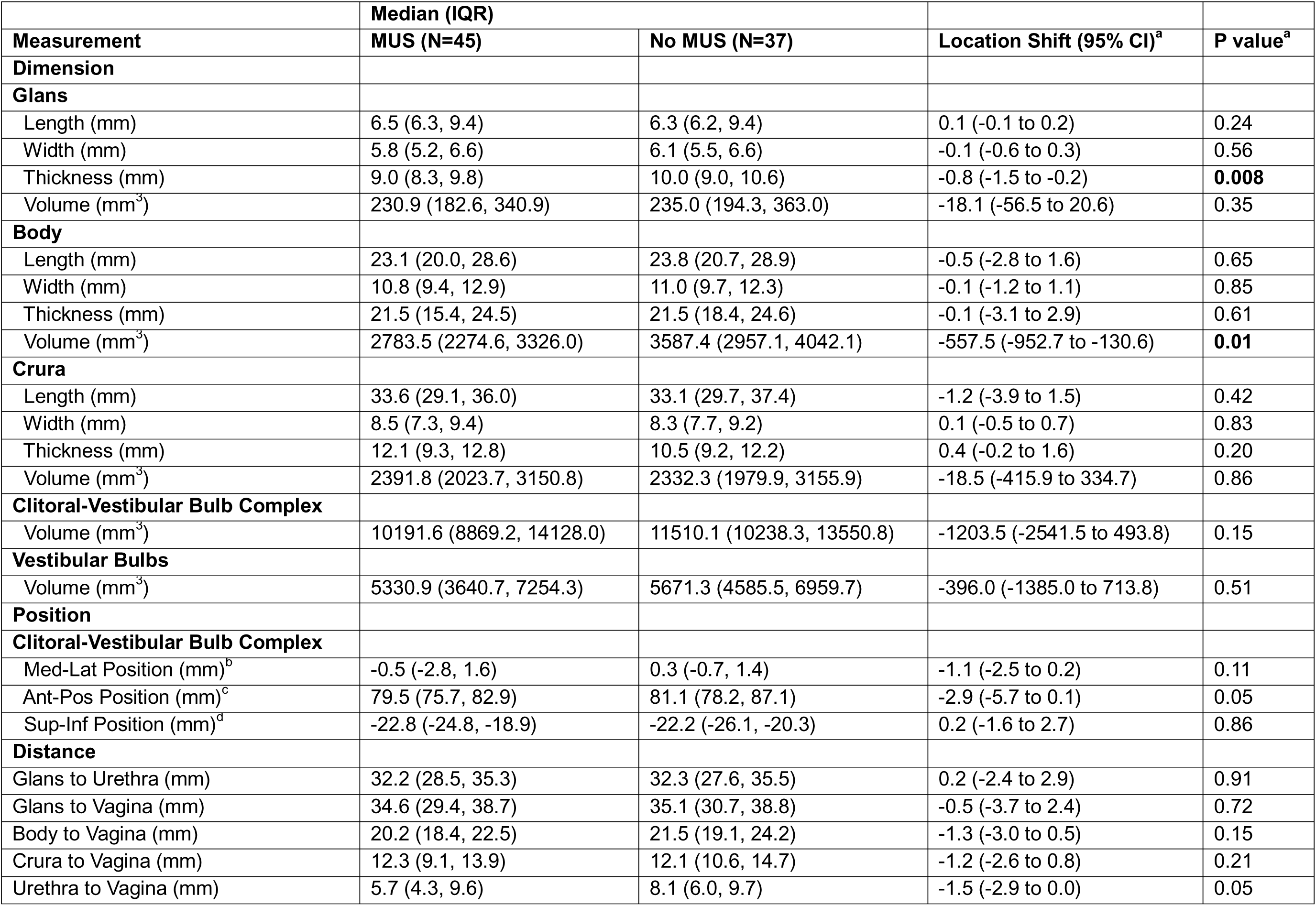
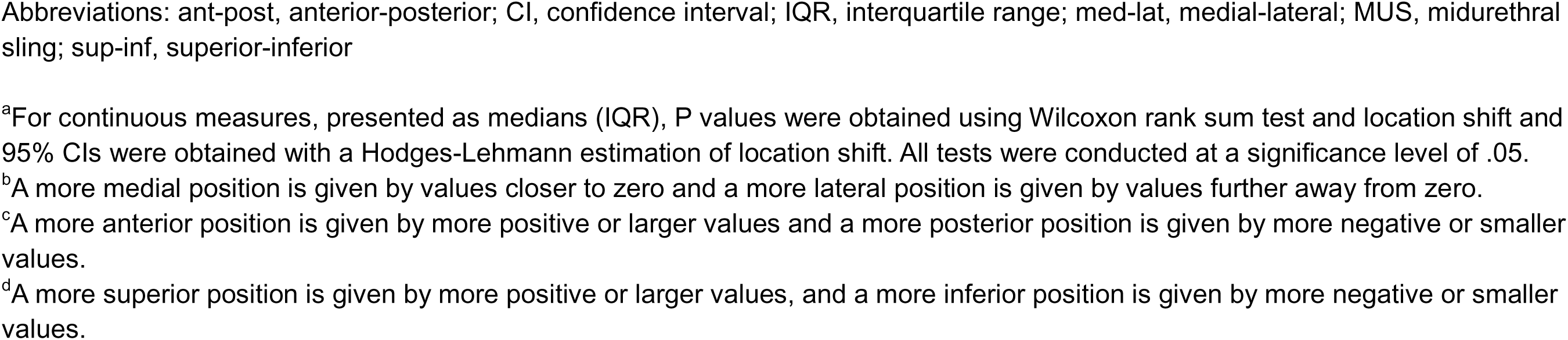
Clitoral-Vestibular Bulb Measurements After Vaginal Prolapse Repair by Concomitant Midurethral Sling (MUS)

|  | **Median (IQR)** |  |  |  |
| --- | --- | --- | --- | --- |
| **Measurement** | **MUS (N=45)** | **No MUS (N=37)** | **Location Shift (95% CI)<sup>a</sup>** | **P value<sup>a</sup>** |
| **Dimension** |  |  |  |  |
| **Glans** |  |  |  |  |
| Length (mm) | 6.5 (6.3, 9.4) | 6.3 (6.2, 9.4) | 0.1 (-0.1 to 0.2) | 0.24 |
| Width (mm) | 5.8 (5.2, 6.6) | 6.1 (5.5, 6.6) | -0.1 (-0.6 to 0.3) | 0.56 |
| Thickness (mm) | 9.0 (8.3, 9.8) | 10.0 (9.0, 10.6) | -0.8 (-1.5 to -0.2) | **0.008** |
| Volume (mm<sup>3</sup>) | 230.9 (182.6, 340.9) | 235.0 (194.3, 363.0) | -18.1 (-56.5 to 20.6) | 0.35 |
| **Body** |  |  |  |  |
| Length (mm) | 23.1 (20.0, 28.6) | 23.8 (20.7, 28.9) | -0.5 (-2.8 to 1.6) | 0.65 |
| Width (mm) | 10.8 (9.4, 12.9) | 11.0 (9.7, 12.3) | -0.1 (-1.2 to 1.1) | 0.85 |
| Thickness (mm) | 21.5 (15.4, 24.5) | 21.5 (18.4, 24.6) | -0.1 (-3.1 to 2.9) | 0.61 |
| Volume (mm<sup>3</sup>) | 2783.5 (2274.6, 3326.0) | 3587.4 (2957.1, 4042.1) | -557.5 (-952.7 to -130.6) | **0.01** |
| **Crura** |  |  |  |  |
| Length (mm) | 33.6 (29.1, 36.0) | 33.1 (29.7, 37.4) | -1.2 (-3.9 to 1.5) | 0.42 |
| Width (mm) | 8.5 (7.3, 9.4) | 8.3 (7.7, 9.2) | 0.1 (-0.5 to 0.7) | 0.83 |
| Thickness (mm) | 12.1 (9.3, 12.8) | 10.5 (9.2, 12.2) | 0.4 (-0.2 to 1.6) | 0.20 |
| Volume (mm<sup>3</sup>) | 2391.8 (2023.7, 3150.8) | 2332.3 (1979.9, 3155.9) | -18.5 (-415.9 to 334.7) | 0.86 |
| **Clitoral-Vestibular Bulb Complex** |  |  |  |  |
| Volume (mm<sup>3</sup>) | 10191.6 (8869.2, 14128.0) | 11510.1 (10238.3, 13550.8) | -1203.5 (-2541.5 to 493.8) | 0.15 |
| **Vestibular Bulbs** |  |  |  |  |
| Volume (mm<sup>3</sup>) | 5330.9 (3640.7, 7254.3) | 5671.3 (4585.5, 6959.7) | -396.0 (-1385.0 to 713.8) | 0.51 |
| **Position** |  |  |  |  |
| **Clitoral-Vestibular Bulb Complex** |  |  |  |  |
| Med-Lat Position (mm)<sup>b</sup> | -0.5 (-2.8, 1.6) | 0.3 (-0.7, 1.4) | -1.1 (-2.5 to 0.2) | 0.11 |
| Ant-Pos Position (mm)<sup>c</sup> | 79.5 (75.7, 82.9) | 81.1 (78.2, 87.1) | -2.9 (-5.7 to 0.1) | 0.05 |
| Sup-Inf Position (mm)<sup>d</sup> | -22.8 (-24.8, -18.9) | -22.2 (-26.1, -20.3) | 0.2 (-1.6 to 2.7) | 0.86 |
| **Distance** |  |  |  |  |
| Glans to Urethra (mm) | 32.2 (28.5, 35.3) | 32.3 (27.6, 35.5) | 0.2 (-2.4 to 2.9) | 0.91 |
| Glans to Vagina (mm) | 34.6 (29.4, 38.7) | 35.1 (30.7, 38.8) | -0.5 (-3.7 to 2.4) | 0.72 |
| Body to Vagina (mm) | 20.2 (18.4, 22.5) | 21.5 (19.1, 24.2) | -1.3 (-3.0 to 0.5) | 0.15 |
| Crura to Vagina (mm) | 12.3 (9.1, 13.9) | 12.1 (10.6, 14.7) | -1.2 (-2.6 to 0.8) | 0.21 |
| Urethra to Vagina (mm) | 5.7 (4.3, 9.6) | 8.1 (6.0, 9.7) | -1.5 (-2.9 to 0.0) | 0.05 |

Abbreviations: ant-post, anterior-posterior; CI, confidence interval; IQR, interquartile range; med-lat, medial-lateral; MUS, midurethral sling; sup-inf, superior-inferior
<sup>a</sup>For continuous measures, presented as medians (IQR), P values were obtained using Wilcoxon rank sum test and location shift and 95% CIs were obtained with a Hodges-Lehmann estimation of location shift. All tests were conducted at a significance level of .05.
<sup>b</sup>A more medial position is given by values closer to zero and a more lateral position is given by values further away from zero.
<sup>c</sup>A more anterior position is given by more positive or larger values and a more posterior position is given by more negative or smaller values.
<sup>d</sup>A more superior position is given by more positive or larger values, and a more inferior position is given by more negative or smaller values.

## DISCUSSION

Among SA women after transvaginal prolapse repair, those with concomitant MUS and those who received hysteropexy mesh had poorer postoperative sexual function outcomes related to arousal and orgasm than women with no sling or hysterectomy, respectively. Across the entire cohort, clitoral morphology did not differ by vaginal surgery approach; however, it did vary by MUS placement, where the MUS group (vs. the No-MUS group) had a smaller clitoral glans (thickness) and body (volume).

Meta-analyses have reported that most women experience stable or improved sexual function after MUS placement due to the resolution of stress incontinence symptoms.^4^ This was demonstrated in this study, where sexual function among SA and NSA women within each MUS and vaginal surgery group remained stable or improved after surgery. However, MUS has also been associated with worsened postoperative sexual function among some women (14%–20%),^5^ mainly in arousal and orgasm outcomes, as observed in this present study. Conversely, unlike this study, previous literature reported no difference in postoperative sexual function between vaginal mesh and native tissue repairs, both having a similar rate of worsened sexual function of 10%.^6^ However, despite prolapse symptom improvements, vaginal prolapse surgery may still limit improvement in orgasm frequency or intensity postoperatively, which we found true among the mesh hysteropexy group vs the hysterectomy group.^21^

Recent studies have shown that MUS (1) changes the spatial relationship between the clitoral-vestibular bulb complex (clitoris, vestibular bulbs), urethra, and vagina, and (2) can alter adjacent neurovascular and glandular tissues (via erosion, transection, fibrosis) that are essential to the sexual response. ^22–28^ Overall, these changes can lead to reduced blood flow to the clitoris (impaired engorgement) and decreased sensation (less sensitivity), which may result in difficulties with sexual arousal and orgasm, pain, and eventually clitoral atrophy (shrinking of the clitoris).^23–32^ This could possibly account for the association between MUS placement, poorer sexual arousal/orgasm, and smaller clitoral glans/body size observed in this study, especially given the close proximity of the MUS arms to the clitoral glans/body and the reported link between smaller clitoral glans size and poorer arousal/orgasm outcomes (**Figure 1**).

### Strengths and Limitations

Study strengths include its prospective design, a well-characterized cohort from a randomized clinical trial, use of validated questionnaires, and its 3D approach. A key limitation is that only half of the SUPeR participants enrolled in DEMAND took part in this study, making the cohort unrepresentative of the entire SUPeR group. Nevertheless, baseline characteristics of women enrolled in DEMAND (vs not enrolled) were similar.^13^ Additional limitations were limited generalizability due to cohort homogeneity, timing discrepancies in collecting some PISQ-IR and postoperative MRI data, missing information on sexual practices, no corrections for multiple comparisons, and the lack of preoperative MRI data to assess direct surgery-related changes in clitoral anatomy and sexual function, which should be addressed by prospective studies.

## CONCLUSIONS

Following vaginal prolapse surgery, SA women with MUS (vs without) and who received hysteropexy (vs hysterectomy) had poorer postoperative sexual function outcomes related to arousal and orgasm intensity. Additionally, clitoral-vestibular bulb morphology differed by MUS, where women with MUS (vs without) had a smaller clitoral glans and body. These results corroborate previous reports of poorer sexual function outcomes related to MUS and vaginal mesh placement during vaginal prolapse surgery. The smaller clitoral dimensions observed with MUS align with surgery-induced pathophysiological mechanisms proposed by prior histologic and cadaveric studies. Prospective, longitudinal studies with more robust sexual function outcomes (i.e., including sexual practices and behaviors) are necessary to determine how MUS and vaginal surgery approaches directly impact clitoral-vestibular anatomy and sexual function.

## Data Availability

Data
- Data available: Yes
- Data types: Deidentified participant data
- How to access data: Data from the DEMAND study will be available from the Eunice Kennedy Shriver National Institute of Child Health and Human Development Data and Specimen Hub (DASH) https://dash.nichd.nih.gov/ after completion of planned DEMAND analyses.
- When available: 12-31-2026
Supporting Documents
- Document types: None
Additional Information
- Who can access the data: researchers whose proposed use of the data has been approved by DASH
- Types of analyses: secondary analysis
- Mechanisms of data availability: Through DASH
- Any additional restrictions: The following disclaimer should be added to all abstracts and manuscripts using the PFDN public use datasets: "Data in this report was collected by the Pelvic Floor Disorders Network. This study was supported by grant funding from the Eunice Kennedy Shriver National Institute of Child Health and Human Development, National Institutes of Health. The content of this report is solely the responsibility of the authors and does not necessarily represent the views of the Pelvic Floor Disorders Network investigators or the NIH."

https://pmc.ncbi.nlm.nih.gov/articles/instance/11822609/bin/jamasurg-e246922-s001.pdf

## AUTHOR CONTRIBUTIONS/DATA ACCESS, RESPONSIBILITY, & ANALYSIS

**Drs. Gantz** and **Bowen,** and **Ms. Sridhar** had full access to all the study data and take responsibility for its integrity and the accuracy of its analysis.

- *Concept and design:* Moalli, Rogers, Corton, Andy, Rardin, Hahn, Weidner, Ellington, Gantz.
- *Acquisition, analysis, or interpretation of data:* All authors.
- *Drafting of the manuscript:* All authors.
- *Critical revision of the manuscript for important intellectual content:* All authors.
- *Statistical Analysis:* Bowen, Sridhar, Gantz.
- *Obtained funding:* Moalli, Weidner, Rardin, Gantz.
- *Administrative, technical, or material support:* Moalli, Mazloomdoost, Sridhar, Gantz.
- *Supervision:* Moalli, Mazloomdoost, Gantz.

## CONFLICTS OF INTEREST/FINANCIAL DISCLOSURES

All authors reported funding from the Eunice Kennedy Shriver National Institute of Child Health and Human Development (NICHD) and the National Institutes of Health Office of Research on Women’s Health, and that Boston Scientific Corporation provided partial support for the SUPeR trial through a research grant to the Pelvic Floor Disorders Network (PFDN) Data Coordinating Center, RTI International.

- **Dr. Moalli** reported receiving funding from NIH/Eunice Kennedy Shriver National Institute of Child Health and Human Development (NICHD) (R01HD083383, R01HD097187, R01HD108666) and the Richard King Mellon Foundation; has a patent for Elastomeric Auxetic Membrane for Urogynecological and Abdominal Implantations.
- **Dr. Rogers** reported receiving royalties from UpToDate and travel and stipend from AUGS for Editor in Chief services for Urogynecology
- **Dr. Rardin** reported receiving research support from the NICHD, Reia LLC, and Foundation for Female Health Awareness.
- **Dr. Hahn** reported receiving research support from General Electric and serving as a consultant to HealthLytix.
- **Dr. Weidner** reported receiving grants from the NIH and Ethicon; served as a consultant for Urocure and Inspire Medical outside the submitted work.
- **No other disclosures were reported.**

## FUNDING/SUPPORT

This study was conducted by the *Eunice Kennedy Shriver* NICHD-sponsored PFDN (grant numbers U10 HD054214, U10 HD041267, U10 HD041261, U10 HD069013, U10 HD069025, U10 HD069010, U10 HD069006, U10 HD054215, U01 HD069031) and the National Institutes of Health (NIH) Office of Research on Women’s Health (ORWH). Partial support for the SUPeR trial was provided by Boston Scientific Corporation through a research grant to the PFDN Data Coordinating Center, RTI International. Research training support was provided by the *National Academies of Sciences, Engineering, and Medicine*’s Ford Foundation Predoctoral Fellowship, the *Massachusetts Institute of Technology (MIT) School of Engineering* Postdoctoral Fellowship Program for Engineering Excellence, and the *Icahn School of Medicine at Mount Sinai (ISMMS)* Blavatnik Family Women’s Health Research Institute. The content is solely the responsibility of the authors and does not necessarily reflect the official views of these funding sources.

## ROLE OF THE FUNDER/SPONSOR

The NICHD project scientist for the PFDN at the time of this study, Dr. Mazloomdoost, was involved in the development of the protocol, management of the study, and preparation, review, and approval of the manuscript. Other NIH employees managed the study’s funding. The Ford Foundation, MIT School of Engineering, and Blavatnik Family Women’s Health Research Institute provided support for research training. Boston Scientific had no role in any aspects of this study.

## DISCLAIMER

The content of this manuscript is solely the responsibility of the authors and does not necessarily represent the official views of the NIH, Ford Foundation, MIT, or ISMMS.

## GROUP INFORMATION

Members of the *Eunice Kennedy Shriver* NICHD Pelvic Floor Disorders Network are listed in

**Supplement 2**.

## DATA SHARING STATEMENT

See **Supplement 3**.

## Notes

### Clinical Trial

NCT01802281

### Clinical Protocols

https://pmc.ncbi.nlm.nih.gov/articles/PMC7917148/

### Author Declarations

Ethics committee/IRB of University of California San Diego gave ethical approval for this work. Ethics committee/IRB of Duke University gave ethical approval for this work. Ethics committee/IRB of University of Alabama at Birmingham gave ethical approval for this work. Ethics committee/IRB of Brown University gave ethical approval for this work. Ethics committee/IRB of University of New Mexico gave ethical approval for this work. Ethics committee/IRB of University of Pennsylvania gave ethical approval for this work. Ethics committee/IRB of University of Pittsburgh gave ethical approval for this work. Ethics committee/IRB of Cleveland Clinic gave ethical approval for this work.

## REFERENCES

1. Lukacz ES, Sridhar A, Chermansky CJ, et al. Sexual Activity and Dyspareunia One Year After Surgical Repair of Pelvic Organ Prolapse. Obstetrics and gynecology. 2020;136(3):492. doi:10.1097/AOG.0000000000003992

2. Dunivan GC, Sussman AL, Jelovsek JE, et al. Gaining the patient perspective on pelvic floor disorders’ surgical adverse events. Am J Obstet Gynecol. 2019;220(2):185.e1–185.e10. doi:10.1016/j.ajog.2018.10.033

3. Antosh DD, Dieter AA, Balk EM, et al. Sexual function after pelvic organ prolapse surgery: a systematic review comparing different approaches to pelvic floor repair. Am J Obstet Gynecol. 2021;225(5):475.e1–475.e19. doi:10.1016/j.ajog.2021.05.042

4. Lai S, Diao T, Zhang W, et al. Sexual Functions in Women With Stress Urinary Incontinence After Mid-Urethral Sling Surgery: A Systematic Review and Meta-Analysis of Prospective Randomized and Non-Randomized Studies. J Sex Med. 2020;17(10):1956–1970. doi:10.1016/j.jsxm.2020.07.003

5. Szell N, Komisaruk B, Goldstein SW, Qu X (Harvey), Shaw M, Goldstein I. A Meta-Analysis Detailing Overall Sexual Function and Orgasmic Function in Women Undergoing Midurethral Sling Surgery for Stress Incontinence. Sex Med. 2017;5(2):e84–e93. doi:10.1016/J.ESXM.2016.12.001

6. Liao SC, Huang WC, Su TH, Lau HH. Changes in Female Sexual Function After Vaginal Mesh Repair Versus Native Tissue Repair for Pelvic Organ Prolapse: A Meta-Analysis of Randomized Controlled Trials. J Sex Med. 2019;16(5):633–639. doi:10.1016/j.jsxm.2019.02.016

7. Lowenstein L, Gruenwald I, Itskovitz-Eldor J, Gartman I, Vardi Y. Is there an association between female urinary incontinence and decreased genital sensation? Neurourol Urodyn. 2011;30(7):1291–1294. doi:10.1002/nau.20988

8. Ram R, Jambhekar K, Glanc P, et al. Meshy business: MRI and ultrasound evaluation of pelvic floor mesh and slings. Abdominal Radiology 2020 46:4. 2020;46(4):1414–1442. doi:10.1007/S00261-020-02404-X

9. Oakley SH, Vaccaro CM, Crisp CC, et al. Clitoral size and location in relation to sexual function using pelvic MRI. Journal of Sexual Medicine. 2014;11(4):1013–1022. doi:10.1111/jsm.12450

10. Bowen ST, Moalli PA, Rogers RG, et al. Postoperative Sexual Function After Vaginal Surgery and Clitoral Size, Position, and Shape. JAMA Surg. 2025;160(4):396–406. doi:10.1001/JAMASURG.2024.6922

11. Abdulcadir J, Botsikas D, Bolmont M, et al. Sexual Anatomy and Function in Women With and Without Genital Mutilation: A Cross-Sectional Study. Journal of Sexual Medicine. 2016;13(2):226–237. doi:10.1016/j.jsxm.2015.12.023

12. Moalli PA, Bowen ST, Abramowitch SD, et al. Methods for the Defining Mechanisms of Anterior Vaginal Wall Descent (DEMAND) Study. Int Urogynecol J. Published online September 1, 2020:1–10. doi:10.1007/s00192-020-04511-1

13. Bowen ST, Moalli PA, Abramowitch SD, et al. Defining mechanisms of recurrence following apical prolapse repair based on imaging criteria. Am J Obstet Gynecol. Published online June 1, 2021. doi:10.1016/J.AJOG.2021.05.041

14. Nager CW, Visco AG, Richter HE, et al. Effect of vaginal mesh hysteropexy vs vaginal hysterectomy with uterosacral ligament suspension on treatment failure in women with uterovaginal prolapse: A randomized clinical trial. JAMA - Journal of the American Medical Association. 2019;322(11):1054–1065. doi:10.1001/jama.2019.12812

15. Rogers RG, Rockwood TH, Constantine ML, et al. A new measure of sexual function in women with pelvic floor disorders (PFD): The Pelvic Organ Prolapse/Incontinence Sexual Questionnaire, IUGA-Revised (PISQ-IR). Int Urogynecol J. 2013;24(7):1091–1103. doi:10.1007/S00192-012-2020-8

16. Constantine ML, Pauls RN, Rogers RR, Rockwood TH. Validation of a single summary score for the Prolapse/Incontinence Sexual Questionnaire–IUGA revised (PISQ-IR). Int Urogynecol J. 2017;28(12):1901–1907. doi:10.1007/S00192-017-3373-9

17. Fedorov A, Beichel R, Kalpathy-Cramer J, et al. 3D Slicer as an image computing platform for the Quantitative Imaging Network. Magn Reson Imaging. 2012;30(9):1323–1341. doi:10.1016/j.mri.2012.05.001

18. Sinex DCE, Bowen ST, Kashkoush A, et al. The establishment of a 3D anatomical coordinate system for defining vaginal axis and spatial position. Comput Methods Programs Biomed. 2021;208:106175. doi:10.1016/J.CMPB.2021.106175

19. Bowen ST, Dutta A, Rytel K, Abramowitch SD, Rogers RG, Moalli PA. 3D quantitative analysis of normal clitoral anatomy in nulliparous women by MRI. Int Urogynecol J. 2022;33(6):1649–1657. doi:10.1007/s00192-022-05172-y

20. Bowen ST, Moalli PA, Abramowitch S, et al. Vaginal morphology and position associated with prolapse recurrence after vaginal surgery: a secondary analysis of the DEMAND study. Authorea Preprints. Published online January 20, 2023. doi:10.22541/AU.167423421.16369342/V1

21. Jafarzade A, Ulu I. Sexual dysfunction in patients after cystocele surgery. Is the g-spot a myth or reality? European Journal of Obstetrics and Gynecology and Reproductive Biology. 2023;290:74–77. doi:10.1016/j.ejogrb.2023.09.009

22. Oakley SH, Mutema GK, Crisp CC, et al. Innervation and Histology of the Clitoral–Urethal Complex: A Cross-Sectional Cadaver Study. J Sex Med. 2013;10(9):2211–2218. doi:10.1111/JSM.12230

23. Giovannetti O, Tomalty D, Gaudet D, et al. Immunohistochemical Investigation of Autonomic and Sensory Innervation of Anterior Vaginal Wall Female Periurethral Tissue: A Study of the Surgical Field of Mid-Urethral Sling Surgery Using Cadaveric Simulation. J Sex Med. 2021;18(7):1167–1180. doi:10.1016/J.JSXM.2021.05.002

24. Achtari C, Mckenzie BJ, Hiscock R, et al. Anatomical study of the obturator foramen and dorsal nerve of the clitoris and their relationship to minimally invasive slings. International Urogynecology Journal 2005 17:4. 2005;17(4):330–334. doi:10.1007/S00192-005-0004-7

25. Rahn DD, Marinis SI, Schaffer JI, Corton MM. Anatomical path of the tension-free vaginal tape: Reassessing current teachings. Am J Obstet Gynecol. 2006;195(6):1809–1813. doi:10.1016/j.ajog.2006.07.009

26. Tomalty D, Giovannetti O, Gaudet D, et al. The prostate in women: an updated histological and immunohistochemical profile of the female periurethral glands and their relationship to an implanted midurethral sling. J Sex Med. 2023;20(5):612–625. doi:10.1093/JSXMED/QDAC046

27. Bekker MD, Hogewoning CRC, Wallner C, Elzevier HW, Deruiter MC. The somatic and autonomic innervation of the clitoris; preliminary evidence of sexual dysfunction after minimally invasive slings. J Sex Med. 2012;9(6):1566–1578. doi:10.1111/J.1743-6109.2012.02711.X

28. Tappy EE, Ramirez D, Stork A, Carrick K, Hamner J, Corton M. Somatic and autonomic nerve distribution of the urethra, periurethral tissue, and anterior vaginal wall: an immunohistochemical study in adult female cadavers. Am J Obstet Gynecol. 2023;228(3):S802–S803. doi:10.1016/j.ajog.2022.12.027

29. Christofferson M, Barnard J, Montoya TI. Clitoral Pain Following Retropubic Midurethral Sling Placement. Sex Med. 2015;3(4):346–348. doi:10.1002/sm2.95

30. Oiwaa Y, Watanabea T, Sadahiraa T, et al. Clitoral Blood Flow Changes after Surgery with Tension-Free Vaginal Mesh for Pelvic Organ Prolapse. Acta Med Okayama. 2019;73(1):21–27. doi:10.18926/AMO/56455

31. Castiglione F, Bergamini A, Albersen M, et al. Pelvic nerve injury negatively impacts female genital blood flow and induces vaginal fibrosis - Implications for human nerve-sparing radical hysterectomy. BJOG. 2015;122(11):1457–1465. doi:10.1111/1471-0528.13506

32. Lowenstein L, Mustafa S, Gartman I, Gruenwald I. Effect of Midurethral Sling Surgery on Vaginal Sensation. J Sex Med. 2016;13(3):389–392. doi:10.1016/j.jsxm.2016.01.006

