## Supplemental Material for "Sexual Function and Clitoral Anatomy after Vaginal Surgery with and without Midurethral Sling"

**Supplement 1:**

**eTable 1.** Inclusion and Exclusion Criteria

**eFigure 1.** Flow of Participants in the Defining Mechanisms of Anterior Vaginal Wall Descent (DEMAND) Ancillary Study by Concomitant Midurethral Sling

**eFigure 2.** Flow of Participants in the Defining Mechanisms of Anterior Vaginal Wall Descent (DEMAND) Ancillary Study by Type of Vaginal Surgery

**eTable 2.** Pelvic Organ Prolapse Quantification (POP-Q) Measurements

**eTable 3.** Baseline and Postoperative Demographic and Clinical Characteristics After Vaginal Prolapse Repair by Type of Vaginal Surgery

**eTable 4.** Baseline and Postoperative Sexual Function Outcomes After Vaginal Prolapse Repair by Type of Vaginal Surgery

**eTable 5.** Clitoral-Vestibular Bulb Measurements After Vaginal Prolapse Repair by Type of Vaginal Surgery

This supplemental material has been provided by the authors to offer readers extra information about their work.

**eTable 1.** Inclusion and Exclusion Criteria

| **Study Enrollment Eligibility** | **Inclusion Criteria** | **Exclusion Criteria** |
| --- | --- | --- |
| DEMAND Primary Study | - Women aged 21 or older who have completed childbearing - Prolapse beyond the hymen (defined as Ba, Bp, or C>0 cm) - Uterine descent into at least the lower half of the vagina [defined as point C>-TVL/2)] - Bothersome bulge symptoms as indicated on question 3 of the PFDI-20 form relating to ‘sensation of bulging’ or ‘something falling out’ - Desires vaginal surgical treatment for uterovaginal prolapse - Available for up to 60-month follow-up - Amenorrhea for the past 12 months from either menopause or endometrial ablation - Not pregnant, not at risk for pregnancy, or agree to contraception if at risk for pregnancy (only applicable to the rare endometrial ablation patient) - Eligible for no cervical cancer screening for at least 3 years | - Previous synthetic material (placed vaginally or abdominally) to augment POP repair - Known previous uterosacral or sacrospinous uterine suspension - Known adverse reaction to synthetic mesh or biological grafts; these complications include but are not limited to erosion, fistula, or abscess - Chronic pelvic pain - Pelvic radiation - Cervical elongation—defined as an expectation that the C point would be Stage 2 or greater postoperatively if a hysteropexy was performed (Note: cervical shortening or trachelectomy is not an allowed intraoperative procedure within the hysteropexy treatment group) - Women at increased risk of cervical dysplasia requiring cervical cancer screening more often than every 3 years [e.g., HIV+ status, immunosuppression because of transplant related medications, Diethylstilbestrol (DES) exposure in utero, or previous treatment for cervical intraepithelial neoplasia (CIN)2, CIN3, or cancer] - Uterine abnormalities (symptomatic uterine fibroids, polyps, endometrial hyperplasia, endometrial cancer, or - any uterine disease that precluded prolapse repair with uterine preservation in the opinion of the surgeon) - Indication for ovarian removal (adnexal mass, BRCA 1/2 positivity, family history of ovarian cancer) - Current condition of amenorrhea caused by exogenous sex steroids or hypothalamic conditions |
| **MRI Analysis Eligibility** | **Inclusion Criteria** | **Exclusion Criteria** |
| DEMAND Primary Study | - N/A | - Failure to capture the entire vagina - MRI taken after reoperation - Incomplete MRI |
| DEMAND Ancillary Study | - N/A | - Poor demarcation of vaginal borders |

**eFigure 1.** Flow of Participants in the Defining Mechanisms of Anterior Vaginal Wall Descent (DEMAND) Ancillary Study by Concomitant Midurethral Sling

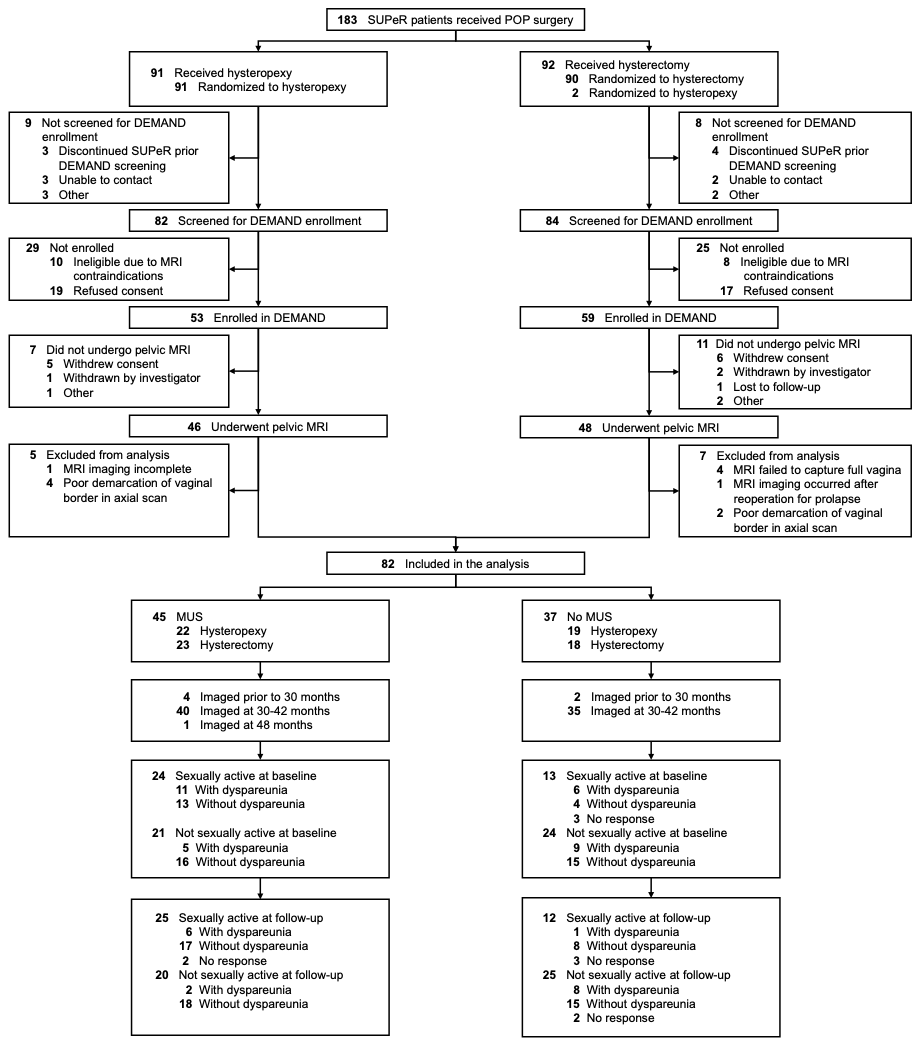

Abbreviations: DEMAND, Defining Mechanisms of Anterior Vaginal Wall Descent; MRI, magnetic resonance imaging; MUS, midurethral sling; POP, pelvic organ prolapse; SUPeR, Study of Uterine Prolapse Procedures-Randomized

Three patients—2 received hysterectomy and 1 received hysteropexy—were found ineligible for the intervention in the parent Study of Uterine Prolapse Procedures-Randomized (SUPeR) trial. Among the women who received hysterectomy, two underwent hysterectomy and sacrospinous ligament suspension.

**eFigure 2.** Flow of Participants in the Defining Mechanisms of Anterior Vaginal Wall Descent (DEMAND) Ancillary Study by Type of Vaginal Surgery

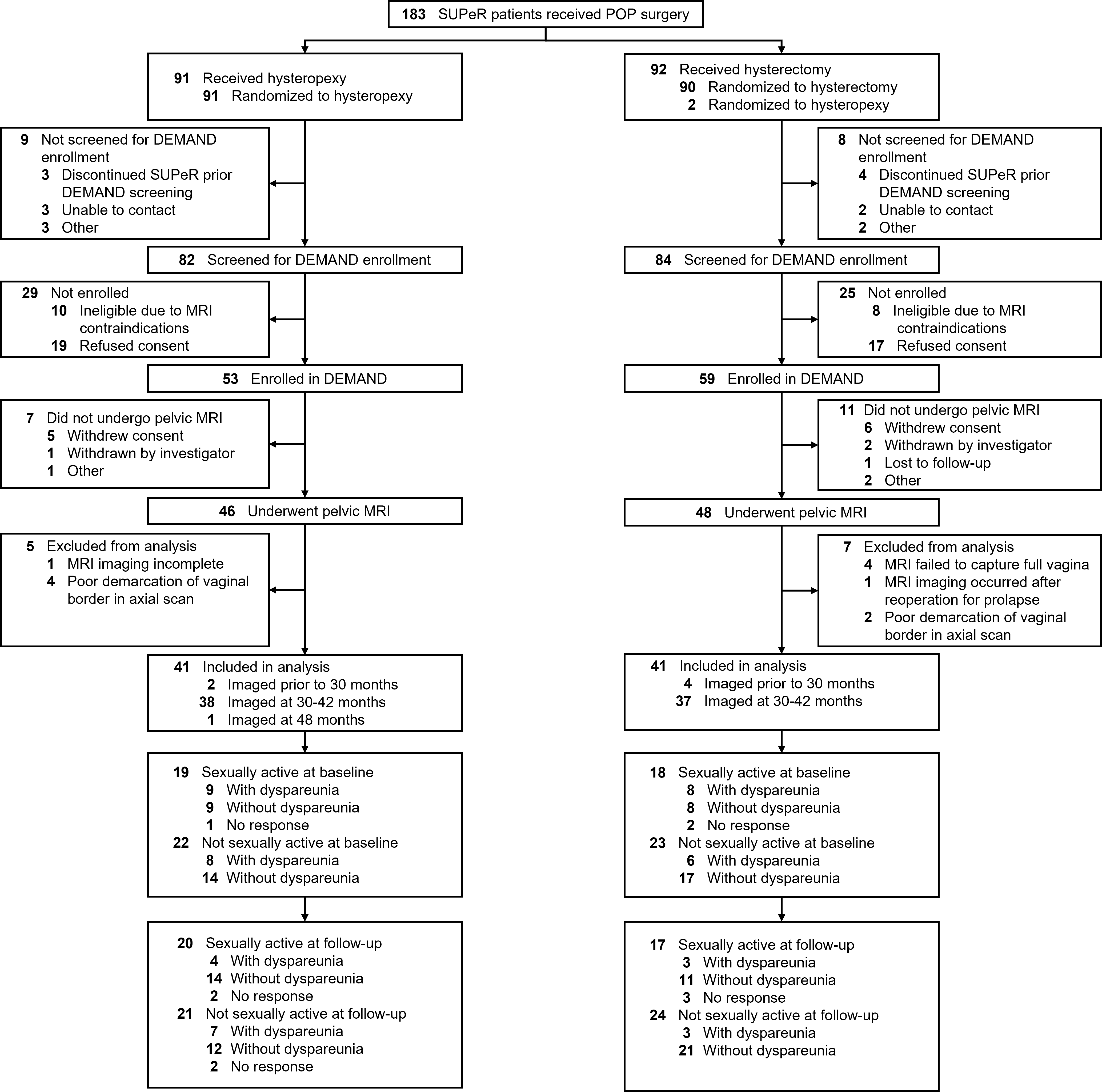

Abbreviations: DEMAND, Defining Mechanisms of Anterior Vaginal Wall Descent; MRI, magnetic resonance imaging; POP, pelvic organ prolapse; SUPeR, Study of Uterine Prolapse Procedures-Randomized

Three patients—2 received hysterectomy and 1 received hysteropexy—were found ineligible for the intervention in the parent Study of Uterine Prolapse Procedures-Randomized (SUPeR) trial. Among the women who received hysterectomy, two underwent hysterectomy and sacrospinous ligament suspension.

**eTable 2.** Pelvic Organ Prolapse Quantification (POP-Q) Measurements

| **POP-Q Measure** | **Description** | **Range** |
| --- | --- | --- |
| **Aa** | Anterior vaginal wall 3 cm proximal to hymen | -3 cm to +3 cm |
| **Ba** | Most dependent part of anterior vaginal wall | -3 cm to +TVL |
| **C** | Cervix or vaginal cuff |  |
| **D** | Posterior fornix |  |
| **Ap** | Posterior vaginal wall 3 cm proximal to hymen | -3 cm to +3 cm |
| **Bp** | Most dependent part of posterior vaginal wall | -3 cm to +TVL |
| **GH** | Genital hiatus, measured from the middle of the external urethral meatus to the posterior border of the hymen | + number |
| **PB** | Perineal body, measured from the posterior border of the hymen to the middle of the anal opening | + number |
| **TVL** | Total vaginal length, measured from the posterior fornix to the hymen when Point C or D is fully reduced to its normal position | + number |

Abbreviations: GH, genital hiatus; PB, perineal body; POP-Q, pelvic organ prolapse quantification; TVL, total vaginal length

**eReference**

Bump, R. C., Mattiasson, A., Bø, K., Brubaker, L. P., DeLancey, J. O., Klarskov, P., ... & Smith, A. R. (1996). The standardization of terminology of female pelvic organ prolapse and pelvic floor dysfunction. *American journal of obstetrics and gynecology*, 175(1), 10-17. doi:10.1016/S0002-9378(96)70243-0

**eTable 3.** Baseline and Postoperative Demographic and Clinical Characteristics After Vaginal Prolapse Repair by Type of Vaginal Surgery

|  |  | **Median (IQR)** | |  |  |
| --- | --- | --- | --- | --- | --- |
| **Characteristic** | **Total No.^a^** | **Hysteropexy (n = 41)** | **Hysterectomy (n = 41)** | **Risk Difference/ Location Shift (95% CI)^b^** | **P value^b^** |
| **Baseline Patient Demographics** |  |  |  |  |  |
| Age, years | 82 | 63.5 (59.7, 68.1) | 65.4 (61.3, 72.3) | -1.6 (-5.5 to 1.5) | 0.34 |
| White | 82 | 33 (81) | 34 (83) | -2 (-20 to 15) | >0.99 |
| Hispanic or Latina, No./total No. (%) | 78 | 5/39 (13) | 4/39 (10) | 3 (-13 to 18) | >0.99 |
| Married/living with partner | 82 | 25 (61) | 27 (66) | -5 (-26 to 16) | 0.82 |
| Higher education after high school, No./total No. (%) | 80 | 26/39 (67) | 29/41 (71) | -4 (-25 to 17) | 0.81 |
| Medicaid/Medicare | 82 | 19 (46) | 21 (51) | -5 (-27 to 18) | 0.83 |
| **Baseline Medical History** |  |  |  |  |  |
| Height (cm) | 82 | 160.0 (155.0, 165.0) | 160.0 (157.0, 165.0) | 0.0 (-3.0 to 3.0) | 0.82 |
| Weight (kg) | 82 | 73.0 (66.0, 81.0) | 70.0 (64.0, 79.0) | 2.0 (-3.0 to 7.0) | 0.48 |
| Body mass index (kg/m^2^)^c^ | 82 | 28.7 (25.4, 31.6) | 26.3 (24.2, 30.1) | 1.2 (-0.8 to 2.9) | 0.21 |
| Obese (BMI ≥ 30) | 82 | 14 (34) | 13 (32) | 2 (-18 to 23) | >0.99 |
| Gravidity | 82 | 3.0 (2.0, 4.0) | 3.0 (2.0, 4.0) | 0.0 (0.0 to 1.0) | 0.37 |
| Cesarean delivery | 82 | 2 (5) | 3 (7) | -2 (-16 to 11) | >0.99 |
| Vaginal parity | 82 | 3.0 (2.0, 3.0) | 2.0 (2.0, 3.0) | 0.0 (0.0 to 1.0) | 0.46 |
| Postmenopausal | 82 | 39 (95) | 41 (100) | -5 (-17 to 4) | 0.49 |
| Estrogen use | 82 | 17 (42) | 14 (34) | 7 (-14 to 28) | 0.65 |
| Current or historical smoker | 82 | 9 (22) | 10 (24) | -2 (-21 to 16) | >0.99 |
| Diabetes | 82 | 5 (12) | 4 (10) | 2 (-13 to 18) | >0.99 |
| Pulmonary disease^d^ | 82 | 4 (10) | 3 (7) | 2 (-11 to 17) | >0.99 |
| Cardiovascular disease^e^ | 82 | 4 (10) | 2 (5) | 5 (-8 to 19) | 0.68 |
| Prior pelvic organ prolapse surgery | 82 | 0 (0) | 3 (7) | -7 (-20 to 2) | 0.24 |
| Prior stress urinary incontinence surgery | 82 | 3 (7) | 2 (5) | 2 (-11 to 16) | >0.99 |
| **Baseline Pelvic Floor Measurements** |  |  |  |  |  |
| POP-Q measurement, cm^f^ |  |  |  |  |  |
| Ba | 82 | 3.0 (1.5, 4.0) | 2.0 (1.0, 4.0) | 0.0 (-1.0 to 1.0) | 0.72 |
| Bp | 82 | 0.0 (-2.0, 2.0) | 0.5 (-1.0, 2.0) | -1.0 (-2.0 to 0.5) | 0.25 |
| C | 82 | -2.0 (-3.0, 1.0) | 0.0 (-2.0, 2.0) | -1.0 (-2.0 to 1.0) | 0.33 |
| GH (strain) | 82 | 4.0 (4.0, 5.0) | 4.0 (4.0, 5.0) | 0.0 (0.0 to 0.5) | 0.65 |
| PB (strain) | 82 | 3.0 (2.5, 3.5) | 3.0 (2.5, 4.0) | 0.0 (-0.5 to 0.5) | 0.97 |
| TVL | 82 | 9.0 (8.0, 10.0) | 9.0 (8.0, 10.0) | 0.0 (0.0 to 1.0) | 0.68 |
| Advanced POP-Q stage (≥3)^g^ | 82 | 32 (78) | 30 (73) | 5 (-14 to 24) | 0.80 |
| Apical prolapse (POP-Q C>0)^h^ | 82 | 17 (42) | 19 (46) | -5 (-26 to 17) | 0.82 |
| Anterior prolapse (POP-Q Aa or Ba >0)^i^ | 82 | 39 (95) | 40 (98) | -2 (-14 to 9) | >0.99 |
| Posterior prolapse (POP-Q Ap or Bp >0)^j^ | 82 | 17 (42) | 21 (51) | -10 (-31 to 12) | 0.51 |
| Any vaginal bulge^k^ | 82 | 41 (100) | 41 (100) |  |  |
| Bothersome vaginal bulge, No./total No. (%)^l^ | 81 | 39/40 (98) | 41/41 (100) | -3 (-13 to 7) | 0.49 |
| **Baseline Concomitant Operative Procedures** |  |  |  |  |  |
| Anterior prolapse repair | 82 | 36 (88) | 31 (76) | 12 (-6 to 29) | 0.25 |
| Posterior prolapse repair/perineorrhaphy | 82 | 24 (59) | 25 (61) | -2 (-24 to 19) | >0.99 |
| Urinary incontinence repair | 82 | 22 (54) | 23 (56) | -2 (-24 to 20) | >0.99 |
| Retropubic Sling | 45 | 16 (73) | 16 (70) | 3 (-24 to 30) | >0.99 |
| Transobturator Sling | 45 | 6 (27) | 7 (30) | -3 (-30 to 24) | >0.99 |
| **Postoperative Pelvic Floor Measurements** |  |  |  |  |  |
| POP-Q measurement, cm^f^ |  |  |  |  |  |
| Ba | 82 | -1.5 (-2.0, 0.0) | -1.0 (-1.0, 0.5) | -1.0 (-1.5 to 0.0) | **0.02** |
| Bp | 82 | -2.0 (-3.0, 0.0) | -2.0 (-3.0, -1.0) | 0.0 (0.0 to 1.0) | 0.59 |
| C | 82 | -6.0 (-7.0, -5.0) | -6.0 (-7.0, -5.0) | 0.0 (-1.0 to 0.0) | 0.42 |
| GH (strain) | 82 | 3.0 (2.5, 4.0) | 3.0 (3.0, 4.0) | 0.0 (-0.5 to 0.0) | 0.60 |
| PB (strain) | 82 | 3.0 (3.0, 4.0) | 4.0 (3.0, 4.0) | 0.0 (-0.5 to 0.5) | 0.91 |
| TVL | 82 | 9.0 (8.0, 9.0) | 8.0 (7.0, 8.5) | 1.0 (0.0 to 1.0) | **0.002** |
| Change from Baseline POP-Q measurement, cm^f^ |  |  |  |  |  |
| Ba | 82 | -4.0 (-5.0, -3.0) | -3.5 (-4.0, -2.0) | -1.0 (-2.0 to 0.0) | 0.08 |
| Bp | 82 | -2.0 (-3.0, 0.0) | -2.0 (-4.0, -0.5) | 1.0 (0.0 to 2.0) | 0.16 |
| C | 82 | -5.0 (-6.5, -3.0) | -6.0 (-8.0, -3.0) | 1.0 (-1.0 to 2.0) | 0.30 |
| GH (strain) | 82 | -1.0 (-2.0, -0.5) | -1.0 (-1.5, -0.5) | 0.0 (-1.0 to 0.0) | 0.38 |
| PB (strain) | 82 | 0.5 (0.0, 1.0) | 0.5 (0.0, 1.0) | 0.0 (0.0 to 0.5) | 0.62 |
| TVL | 82 | -0.5 (-1.0, 0.0) | -1.0 (-2.0, 0.0) | 1.0 (0.0 to 1.0) | **0.009** |
| Apical prolapse (POP-Q C>0)^h^ | 82 | 2 (5) | 1 (2) | 2 (-9 to 14) | >0.99 |
| Anterior prolapse (POP-Q Aa or Ba >0)^i^ | 82 | 4 (10) | 11 (27) | -17 (-34 to 0) | 0.08 |
| Posterior prolapse (POP-Q Ap or Bp >0)^j^ | 82 | 4 (10) | 3 (7) | 2 (-11 to 17) | >0.99 |
| Any vaginal bulge^k^ | 82 | 4 (10) | 5 (12) | -2 (-18 to 13) | >0.99 |
| Bothersome vaginal bulge, No./total No. (%)^l^ | 82 | 4 (10) | 2 (5) | 5 (-8 to 19) | 0.68 |

Abbreviations: BMI, body mass index; GH, genital hiatus; MRI, resonance imaging; MUS, midurethral sling; PB, perineal body; PFDI 20, Pelvic Floor Distress Inventory 20; POP-Q, Pelvic Organ Prolapse Quantification; SUPeR, Study of Uterine Prolapse Procedures–Randomized; TVL, total vaginal length.

^a^A total number less than 82 indicates missing participant data.

^b^For nominal categorical measures, presented as counts (percentages), the P values were obtained from Fisher exact test and exact risk difference and 95%CI limits were obtained by exact methods based on the score statistic. For continuous measures, presented as medians (IQR), P values were obtained using Wilcoxon rank sum test and location shift and 95% CIs were obtained with a Hodges-Lehmann estimation of location shift. All tests were conducted at a significance level of .05.

^c^Calculated as weight in kilograms divided by height in meters squared.

^d^Pulmonary disease includes any of the following: asthma, chronic obstructive pulmonary disease, acute respiratory distress syndrome, and emphysema.

^e^Cardiovascular disease includes any of the following: angina, congenital heart failure/heart disease, heart attack, stroke/transient ischemic attack, and peripheral vascular disease.

^f^POP-Q point Ba is the most distal position of the upper anterior vaginal wall. POP-Q point Bp is the most distal position of the upper posterior vaginal wall. POP-Q point C is the most distal edge of the cervix or leading edge of the vaginal cuff (hysterectomy scar). POP-Q GH is measured from the middle of the external urethral meatus to the posterior border of the hymen. POP-Q PB is measured from the posterior border of the hymen to the middle of the anal opening. POP-Q TVL is measured from the posterior fornix to the hymen when point C or D is fully reduced to its normal position. **eTable 2** in Supplement 1 contains further details.

^g^POP-Q stages: stage 2, the vagina is prolapsed between 1 cm above the hymen and 1 cm below the hymen; stage 3, the vagina is prolapsed more than 1 cm beyond the hymen but is not everted within 2 cm of its length; stage 4, the vagina is everted to within 2 cm of its length.

^h^POP-Q point C is the most distal edge of the cervix or leading edge of the vaginal cuff (hysterectomy scar). **eTable 2** in Supplement 1 contains further details.

^i^POP-Q point Aa is the midline of the anterior vaginal wall 3 cm proximal to the external urethral meatus. POP-Q point Ba is the most distal position of the upper anterior vaginal wall. **eTable 2** in Supplement 1 contains further details.

^j^POP-Q point Ap is the midline of the posterior vaginal wall 3 cm proximal to the hymen. POP-Q point Bp is the most distal position of the upper posterior vaginal wall. **eTable 2** in Supplement 1 contains further details.

^k^Any vaginal bulge symptoms is defined as a positive response to item 3 on the PFDI 20, “Do you usually have a bulge or something falling out that you can see or feel in the vaginal area?”

^l^Bothersome vaginal bulge symptoms is defined as a positive response to any vaginal bulge symptoms and a response of somewhat,” “moderately,” or “quite a bit” to the follow-up question “How much does this bother you?” for item 3 on the PFDI 20.

**eTable 4.** Baseline and Postoperative Sexual Function Outcomes After Vaginal Prolapse Repair by Type of Vaginal Surgery

|  |  | **Median (IQR)** | |  |  |
| --- | --- | --- | --- | --- | --- |
| **Baseline Patient-Reported Sexual Function Outcomes** | **Total No.** | **Hysteropexy (n = 41)** | **Hysterectomy (n = 41)** | **Risk Difference/ Location Shift (95% CI)^a^** | ***P* Value^a^** |
| Sexually active^b^ | 82 | 19 (46) | 18 (44) | 2 (-20 to 24) | >0.99 |
| **Among Sexually Active before Vaginal Prolapse Repair^b^** |  | **N = 19** | **N = 18** |  |  |
| Dyspareunia, No./total No. (%)^c^ | 34 | 9/18 (50) | 8/16 (50) | 0 (-34 to 34) | >0.99 |
| PISQ-IR condition impact score^d^ | 37 | 3.0 (2.5, 4.0) | 3.0 (2.5, 4.0) | 0.0 (-0.5 to 0.5) | 0.99 |
| PISQ-IR condition specific score^d^ | 37 | 4.7 (4.0, 5.0) | 4.8 (4.3, 5.0) | 0.0 (-0.3 to 0.0) | 0.29 |
| PISQ-IR arousal, orgasm score^d^ | 37 | 3.5 (2.8, 4.0) | 3.3 (2.8, 3.8) | 0.3 (-0.3 to 0.8) | 0.32 |
| PISQ-IR summary score^d^ | 37 | 3.5 (2.9, 4.0) | 3.5 (3.1, 3.8) | 0.0 (-0.4 to 0.5) | 0.84 |
| **Among Not Sexually Active before Vaginal Prolapse Repair^b^** |  | **N = 22** | **N = 23** |  |  |
| Dyspareunia, No./total No. (%)^c^ | 45 | 8 (36) | 6 (26) | 10 (-18 to 37) | 0.53 |
| PISQ-IR condition impact score^d^ | 45 | 1.5 (1.0, 3.0) | 1.3 (1.0, 2.3) | 0.0 (-0.3 to 0.7) | 0.61 |
| PISQ-IR condition specific score^d^ | 45 | 1.7 (1.0, 2.7) | 1.7 (1.0, 2.7) | 0.0 (-0.3 to 0.7) | 0.69 |
| **Postoperative Patient-Reported Sexual Function Outcomes** |  |  |  |  |  |
| Sexually active^b^ | 82 | 20 (49) | 17 (42) | 7 (-15 to 29) | 0.66 |
| **Among Sexually Active after Vaginal Prolapse Repair^b^** |  | **N = 20** | **N = 17** |  |  |
| Dyspareunia, No./total No. (%)^c^ | 32 | 4/18 (22) | 3/14 (21) | 1 (-33 to 31) | >0.99 |
| De novo dyspareunia, No./total No. (%)^e^ | 31 | 1/17 (6) | 0/14 (0) | 6 (-18 to 29) | >0.99 |
| PISQ-IR condition impact score^d^ | 37 | 4.0 (3.5, 4.0) | 4.0 (3.5, 4.0) | 0.0 (0.0 to 0.0) | 0.61 |
| PISQ-IR condition specific score^d^ | 36 | 5.0 (4.7, 5.0) | 5.0 (5.0, 5.0) | 0.0 (0.0 to 0.0) | 0.60 |
| PISQ-IR arousal, orgasm score^d^ | 36 | 3.8 (3.3, 4.0) | 3.8 (3.5, 4.3) | -0.3 (-0.5 to 0.3) | 0.34 |
| PISQ-IR summary score^d^ | 36 | 3.8 (3.3, 4.0) | 3.9 (3.4, 4.1) | -0.1 (-0.4 to 0.2) | 0.40 |
| **Among Not Sexually Active after Vaginal Prolapse Repair^b^** |  | **N = 21** | **N = 24** |  |  |
| Dyspareunia, No./total No. (%)^c^ | 43 | 7/19 (37) | 3/24 (13) | 24 (-2 to 51) | 0.08 |
| De novo dyspareunia, No./total No. (%)^e^ | 43 | 1/19 (5) | 2/24 (8) | -3 (-22 to 18) | >0.99 |
| PISQ-IR condition impact score^d^ | 43 | 1.0 (1.0, 2.3) | 1.0 (1.0, 1.2) | 0.0 (0.0 to 0.0) | 0.31 |
| PISQ-IR condition specific score^d^ | 43 | 1.0 (1.0, 3.0) | 1.0 (1.0, 1.8) | 0.0 (0.0 to 1.3) | 0.18 |
| **Change from Baseline Among Sexually Active^b^** |  | **N = 20** | **N = 17** |  |  |
| PISQ-IR condition impact score^d^ | 32 | 0.0 (0.0, 1.0) | 0.0 (0.0, 1.0) | 0.0 (-0.3 to 0.8) | 0.82 |
| PISQ-IR condition specific score^d^ | 31 | 0.0 (0.0, 0.7) | 0.0 (0.0, 0.7) | 0.0 (-0.3 to 0.3) | 0.93 |
| PISQ-IR arousal, orgasm score^d^ | 31 | 0.0 (-0.1, 0.3) | 0.3 (0.3, 1.0) | -0.5 (-0.8 to 0.0) | **0.01** |
| PISQ-IR summary score^d^ | 31 | 0.2 (-0.1, 0.6) | 0.2 (0.0, 0.4) | -0.1 (-0.4 to 0.3) | 0.56 |
| **Change from Baseline Among Not Sexually Active^b^** |  | **N = 21** | **N = 24** |  |  |
| PISQ-IR condition impact score^d^ | 38 | 0.0 (-1.3, 0.0) | 0.0 (-1.0, 0.0) | 0.0 (-0.7 to 0.7) | 0.92 |
| PISQ-IR condition specific score^d^ | 38 | 0.0 (-1.0, 0.0) | 0.0 (-0.7, 0.0) | 0.0 (-0.3 to 0.7) | 0.83 |

Abbreviations: CI, confidence interval; IQR, interquartile range; MRI, magnetic resonance imaging; PISQ-IR, Pelvic Organ Prolapse-Incontinence Sexual Function Questionnaire-IUGA Revised, SUPeR, Study of Uterine Prolapse Procedures-Randomized.

^a^For nominal categorical measures, presented as counts (percentages), the p-values were obtained from Fishers exact test and exact risk difference and 95% CI limits were obtained by exact methods based on the score statistic. For continuous measures, presented as medians (25th, 75th percentiles), p-values were obtained using Wilcoxon Rank-Sum test and location shift and 95% confidence intervals were obtained with a Hodges-Lehmann estimation of location shift. All tests were conducted at a significance level of .05.

^b^Sexual activity is based on Item 1 of the PISQ-IR which asked whether participants were (1) sexually active with or without a partner or (2) not sexually active at all.

^c^Dyspareunia among sexually active participants is based on Item 11 of the PISQ-IR and is defined as experiencing pain (sometimes, usually, or always have) during sexual intercourse. Dyspareunia among not sexually active participants is based on Item 2e of the PISQ-IR and is defined as not engaging in sexual intercourse due to pain (strongly agree or somewhat agree) during sexual intercourse.

^d^Higher PISQ-IR scores among sexually active women indicate better sexual function. For not sexually active women, higher PISQ-IR scores indicated greater impact on sexual inactivity (i.e., poorer sexual function).

^e^De novo dyspareunia is defined as the absence of dyspareunia at baseline and the prevalence of dyspareunia at SUPeR Visit closest to MRI Imaging.

**eTable 5.** Clitoral-Vestibular Bulb Measurements After Vaginal Prolapse Repair by Type of Vaginal Surgery

|  | **Median (IQR)** | |  |  |
| --- | --- | --- | --- | --- |
| **Measurement** | **Hysteropexy (n = 41)** | **Hysterectomy (n = 41)** | **Location Shift (95% CI)^a^** | **P value^a^** |
| **Dimension** |  |  |  |  |
| **Glans** |  |  |  |  |
| Length (mm) | 6.4 (6.3, 9.4) | 6.4 (6.3, 9.4) | 0.0 (-0.2 to 0.1) | 0.75 |
| Width (mm) | 5.8 (5.3, 6.6) | 6.0 (5.4, 6.6) | -0.1 (-0.6 to 0.3) | 0.46 |
| Thickness (mm) | 9.3 (8.5, 10.0) | 9.7 (8.5, 10.6) | -0.4 (-1.0 to 0.3) | 0.28 |
| Volume (mm^3^) | 227.6 (190.2, 284.2) | 270.0 (186.5, 353.1) | -14.4 (-72.0 to 29.5) | 0.48 |
| **Body** |  |  |  |  |
| Length (mm) | 23.6 (20.0, 28.7) | 23.4 (20.9, 28.7) | -0.2 (-2.5 to 2.0) | 0.85 |
| Width (mm) | 11.0 (9.6, 12.5) | 11.0 (9.4, 12.8) | 0.2 (-0.8 to 1.3) | 0.79 |
| Thickness (mm) | 21.5 (18.3, 24.5) | 21.5 (18.3, 24.6) | -0.1 (-3.0 to 2.9) | 0.78 |
| Volume (mm^3^) | 3138.8 (2508.3, 3866.8) | 3034.5 (2348.3, 3766.0) | 9.1 (-455.3 to 424.6) | 0.96 |
| **Crura** |  |  |  |  |
| Length (mm) | 33.1 (29.5, 36.0) | 33.9 (29.4, 37.7) | -1.0 (-3.6 to 1.7) | 0.44 |
| Width (mm) | 8.3 (7.4, 9.2) | 8.5 (7.7, 9.3) | -0.1 (-0.8 to 0.5) | 0.68 |
| Thickness (mm) | 10.5 (9.3, 12.6) | 12.0 (9.2, 12.4) | 0.1 (-0.7 to 0.6) | 0.89 |
| Volume (mm^3^) | 2391.8 (1961.4, 3150.8) | 2332.3 (2054.1, 3151.2) | 35.4 (-349.5 to 409.8) | 0.84 |
| **Clitoral-Vestibular Bulb Complex** |  |  |  |  |
| Volume (mm^3^) | 10865.9 (9063.4, 13364.3) | 11250.4 (9158.4, 14239.8) | -520.8 (-1994.1 to 1085.6) | 0.48 |
| **Vestibular Bulbs** |  |  |  |  |
| Volume (mm^3^) | 5108.7 (3789.6, 6959.7) | 5515.0 (4394.9, 7778.4) | -530.7 (-1530.3 to 482.5) | 0.33 |
| **Position** |  |  |  |  |
| **Clitoral-Vestibular Bulb Complex** |  |  |  |  |
| Med-Lat Position (mm)^b^ | 0.4 (-2.2, 1.5) | -0.5 (-2.4, 1.4) | 0.5 (-0.8 to 1.7) | 0.35 |
| Ant-Pos Position (mm)^c^ | 79.0 (75.7, 85.2) | 81.5 (78.2, 84.6) | -1.1 (-4.1 to 1.6) | 0.38 |
| Sup-Inf Position (mm)^d^ | -22.0 (-26.8, -19.2) | -22.8 (-24.2, -18.9) | -0.1 (-2.6 to 1.8) | 0.89 |
| **Distance** |  |  |  |  |
| Glans to Urethra (mm) | 32.3 (28.2, 35.5) | 31.5 (27.6, 35.4) | -0.1 (-2.7 to 2.6) | 0.90 |
| Glans to Vagina (mm) | 34.6 (30.5, 37.6) | 35.1 (29.3, 39.0) | -0.9 (-4.0 to 2.0) | 0.54 |
| Body to Vagina (mm) | 19.8 (18.4, 23.6) | 21.1 (19.1, 23.0) | -0.7 (-2.4 to 1.0) | 0.37 |
| Crura to Vagina (mm) | 12.5 (10.1, 13.9) | 12.1 (9.7, 15.4) | 0.3 (-1.4 to 1.9) | 0.71 |
| Urethra to Vagina (mm) | 6.7 (4.5, 9.3) | 6.3 (4.7, 9.7) | -0.3 (-1.7 to 1.2) | 0.70 |

Abbreviations: ant-post, anterior-posterior; CI, confidence interval; IQR, interquartile range; med-lat, medial-lateral; sup-inf, superior-inferior

^a^For continuous measures, presented as medians (IQR), P values were obtained using Wilcoxon rank sum test and location shift and 95% CIs were obtained with a Hodges-Lehmann estimation of location shift. All tests were conducted at a significance level of .05.

^b^A more medial position is given by values closer to zero and a more lateral position is given by values further away from zero.

^c^A more anterior position is given by more positive or larger values and a more posterior position is given by more negative or smaller values.

^d^A more superior position is given by more positive or larger values and a more inferior position is given by more negative or smaller values.

**Supplement 2:** Nonauthor Collaborators (Eunice Kennedy Shriver NICHD Pelvic Floor Disorders Network)

| ***First Name and Middle Initial(s)** | ***Last Name** | ***Suffix (e.g., Jr, III)** | **Academic Degrees** | **Institution** | **Location (city, state/province, country)** | **Role or Contribution, eg, chair, principal investigator** | **Group (if more than 1 Group listed in the byline) and/or Subgroup (e.g., Steering Committee)** |
| --- | --- | --- | --- | --- | --- | --- | --- |
| Kimberly | Ferrante |  | MD, MAS | University of California San Diego Health | San Diego, CA, USA | Study Clinician | PFDN Site |
| Sherella | Johnson |  | N/A | University of California San Diego Health | San Diego, CA, USA | Coordinator | PFDN Site |
| Emily S. | Lukacz |  | MD | University of California San Diego Health | San Diego, CA, USA | Principal Investigator | PFDN Site |
| Charles W. | Nager |  | MD | University of California San Diego Health | San Diego, CA, USA | Study Clinician | PFDN Site |
| Gouri B. | Diwadkar |  | MD | Kaiser Permanente | San Diego, CA, USA | Study Clinician | PFDN Site |
| Keisha Y. | Dyer |  | MD, MPH | Kaiser Permanente | San Diego, CA, USA | Study Clinician | PFDN Site |
| Linda M. | Mackinnon |  | BA, MPH | Kaiser Permanente | San Diego, CA, USA | Coordinator | PFDN Site |
| Jasmine | Tan-Kim |  | MD | Kaiser Permanente | San Diego, CA, USA | Study Clinician | PFDN Site |
| Gisselle | Zazueta-Damian |  | N/A | Kaiser Permanente | San Diego, CA, USA | Coordinator | PFDN Site |
| Cindy | Amundsen |  | MD | Duke University Medical Center | Durham, NC, USA | Study Clinician | PFDN Site |
| Yasmeen | Bruton |  | MA | Duke University Medical Center | Durham, NC, USA | Coordinator | PFDN Site |
| Notorious | Coleman-Taylor |  | MS,MSCR | Duke University Medical Center | Durham, NC, USA | Coordinator | PFDN Site |
| Amie | Kawasaki |  | MD, FACOG | Duke University Medical Center | Durham, NC, USA | Study Clinician | PFDN Site |
| Nicole | Longoria |  | PA-C | Duke University Medical Center | Durham, NC, USA | Coordinator | PFDN Site |
| Shantae | McLean |  | MPH | Duke University Medical Center | Durham, NC, USA | Coordinator | PFDN Site |
| Nazema | Siddiqui |  | MD, MHSc | Duke University Medical Center | Durham, NC, USA | Principal Investigator | PFDN Site |
| Kathy | Carter |  | BSN | University of Alabama at Birmingham, Dept. OB/GYN | Birmingham, AL, USA | Coordinator | PFDN Site |
| Mark E. | Lockhart |  | MD, MPH | University of Alabama at Birmingham, Dept. OB/GYN | Birmingham, AL, USA | Study Clinician | PFDN Site |
| Sunita | Patel |  | N/A | University of Alabama at Birmingham, Dept. OB/GYN | Birmingham, AL, USA | Coordinator | PFDN Site |
| Holly E. | Richter |  | MD, PHD | University of Alabama at Birmingham, Dept. OB/GYN | Birmingham, AL, USA | Principal Investigator | PFDN Site |
| Nancy | Saxon |  | RN | University of Alabama at Birmingham, Dept. OB/GYN | Birmingham, AL, USA | Coordinator | PFDN Site |
| Velria B. | Willis |  | RN, BSN, CCRC | University of Alabama at Birmingham, Dept. OB/GYN | Birmingham, AL, USA | Coordinator | PFDN Site |
| Cassandra | Carberry |  | MD | Alpert Medical School of Brown University | Providence, RI, USA | Study Clinician | PFDN Site |
| Brittany S. | Hampton |  | MD, FACOG | Alpert Medical School of Brown University | Providence, RI, USA | Study Clinician | PFDN Site |
| Nicole | Korbly |  | MD | Alpert Medical School of Brown University | Providence, RI, USA | Study Clinician | PFDN Site |
| Ann S. | Meers |  | BSN, RN, CCRC | Alpert Medical School of Brown University | Providence, RI, USA | Coordinator | PFDN Site |
| Deborah L. | Myers |  | MD | Alpert Medical School of Brown University | Providence, RI, USA | Study Clinician | PFDN Site |
| Vivian W. | Sung |  | MD, MPH | Alpert Medical School of Brown University | Providence, RI, USA | Principal Investigator | PFDN Site |
| Kyle | Wohlrab |  | MD | Alpert Medical School of Brown University | Providence, RI, USA | Study Clinician | PFDN Site |
| Gena | Dunivan |  | MD | University of New Mexico | Albuquerque, NM, USA | Study Clinician | PFDN Site |
| Yuko | Komesu |  | MD | University of New Mexico | Albuquerque, NM, USA | Study Clinician | PFDN Site |
| Peter | Jeppson |  | MD, FACOG, FACS | University of New Mexico | Albuquerque, NM, USA | Study Clinician | PFDN Site |
| Lily | Arya |  | MD, MS | University of Pennsylvania | Philadelphia, PA, USA | Principal Investigator | PFDN Site |
| Lorraine | Flick |  | MS | University of Pennsylvania | Philadelphia, PA, USA | Coordinator | PFDN Site |
| Heidi | Harvie |  | MD, MBA, MSCE | University of Pennsylvania | Philadelphia, PA, USA | Study Clinician | PFDN Site |
| Michelle | Kinglee |  | N/A | University of Pennsylvania | Philadelphia, PA, USA | Coordinator | PFDN Site |
| Ariana | Smith |  | MD | University of Pennsylvania | Philadelphia, PA, USA | Study Clinician | PFDN Site |
| Steven D. | Abramowitch |  | PhD | Magee-Women’s Hospital, Dept. of OB/GYN & Reproductive Sciences | Pittsburgh, PA, USA | Study Engineer | PFDN Site |
| Michael | Bonidie |  | MD | Magee-Women’s Hospital, Dept. of OB/GYN & Reproductive Sciences | Pittsburgh, PA, USA | Study Clinician | PFDN Site |
| Judy | Gruss |  | BSN | Magee-Women’s Hospital, Dept. of OB/GYN & Reproductive Sciences | Pittsburgh, PA, USA | Coordinator | PFDN Site |
| Jonathan | Shepherd |  | MA | Magee-Women’s Hospital, Dept. of OB/GYN & Reproductive Sciences | Pittsburgh, PA, USA | Coordinator | PFDN Site |
| Gary | Sutkin |  | MD | Magee-Women’s Hospital, Dept. of OB/GYN & Reproductive Sciences | Pittsburgh, PA, USA | Study Clinician | PFDN Site |
| Halina M. | Zyczynski |  | MD | Magee-Women’s Hospital, Dept. of OB/GYN & Reproductive Sciences | Pittsburgh, PA, USA | Principal Investigator | PFDN Site |
| Matthew | Barber |  | MD, MHSc | Cleveland Clinic Foundation | Cleveland, OH, USA | Principal Investigator | PFDN Site |
| Annette | Graham |  | RN, BSN | Cleveland Clinic Foundation | Cleveland, OH, USA | Coordinator | PFDN Site |
| Marie Fidela R. | Paraiso |  | MD | Cleveland Clinic Foundation | Cleveland, OH, USA | Study Clinician | PFDN Site |
| Cecile | Ferrando |  | MD | Cleveland Clinic Foundation | Cleveland, OH, USA | Study Clinician | PFDN Site |
| Kate | Burdekin |  | MPH | RTI International | Research Triangle Park, NC, USA | Data Manager | PFDN Data Coordinating Center |
| Michael | Ham |  | BS | RTI International | Research Triangle Park, NC, USA | Biostatistician | PFDN Data Coordinating Center |
| Amanda | Shaffer |  | MSN | RTI International | Research Triangle Park, NC, USA | Coordinator | PFDN Data Coordinating Center |
| Dennis | Wallace |  | PHD | RTI International | Research Triangle Park, NC, USA | Principal Investigator | PFDN Data Coordinating Center |
| Ryan | Whitworth |  | PHD | RTI International | Research Triangle Park, NC, USA | Biostatistician | PFDN Data Coordinating Center |
| Taylor | Swankie |  | MPH | RTI International | Research Triangle Park, NC, USA | Coordinator | PFDN Data Coordinating Center |

**Supplement 3.** Data Sharing Statement

**Data**

- ***Data available:*** Yes
- ***Data types:*** Deidentified participant data
- ***How to access data:*** Data from the DEMAND study will be available from the Eunice Kennedy Shriver National Institute of Child Health and Human Development Data and Specimen Hub (DASH) <https://dash.nichd.nih.gov/> after completion of planned DEMAND analyses.
- ***When available:*** 12-31-2026

**Supporting Documents**

- ***Document types:*** None

**Additional Information**

- ***Who can access the data:*** researchers whose proposed use of the data has been approved by DASH
- ***Types of analyses:*** secondary analysis
- ***Mechanisms of data availability:*** Through DASH
- ***Any additional restrictions:*** The following disclaimer should be added to all abstracts and manuscripts using the PFDN public use datasets:
  - *"Data in this report was collected by the Pelvic Floor Disorders Network. This study was supported by grant funding from the Eunice Kennedy Shriver National Institute of Child Health and Human Development, National Institutes of Health. The content of this report is solely the responsibility of the authors and does not necessarily represent the views of the Pelvic Floor Disorders Network investigators or the NIH."*
